# Urinary Biomarkers for Lung Cancer Detection

**DOI:** 10.1101/2024.07.30.24311186

**Authors:** Alexandre Matov

## Abstract

**Introduction:** The current healthcare system relies largely on a passive approach toward disease detection, which typically involves patients presenting a “chief complaint” linked to a particular set of symptoms for diagnosis. Since all degenerative diseases occur slowly and initiate as changes in the regulation of individual cells within our organs and tissues, it is inevitable that with the current approach to medical care we are bound to discover some illnesses at a point in time when the damage is irreversible and meaningful treatments are no longer available.

**Methods:** There exist organ-specific sets (or panels) of nucleic acids, such as microRNAs (miRs), which regulate and help to ensure the proper function of each of our organs and tissues. Thus, dynamic readout of their relative abundance can serve as a means to facilitate real-time health monitoring. With the advent and mass utilization of next-generation sequencing (NGS), such a proactive approach is currently feasible. Because of the computational complexity of customized analyses of “big data”, dedicated efforts to extract reliable information from longitudinal datasets is key to successful early detection of disease.

**Results:** Here, we present our preliminary results for the analysis of healthy donor samples and drug-naïve lung cancer patients’ samples, for which we identify urinary biomarkers demonstrating that small RNAs can pass through the filtration by the kidneys.

**Conclusions:** We provide a proof-of-principle that it is possible to perform non-invasive health monitoring by sequencing of urinary small RNAs and that traces of neoplastic transformation originating in organs that are not adjacent to the urinary tract, like the lungs, can also be detected in urine.

## INTRODUCTION

The cancer incidences and mortality rates worldwide demonstrate that for some cancers, such as lung, stomach, liver, esophagus, leukemia and pancreas cancer, the diagnosis is almost inevitably linked to a loss of life. The 5-years survival rate for patients with lung cancer, the “biggest killer”, is 47% for stage I and can be as low as 2% for stage IV (Fig. 1), which underscores the importance of early detection (WHO, 2017). It will be beneficial to develop molecular diagnostics tests based on nucleic acid markers for monitoring of cardiovascular, neoplastic, and diseases of the nervous system based on the longitudinal analyses of body fluids. In the context of the analysis of samples from healthy individuals, the significance of such approaches will be in providing novel ways for detection of disease. More specifically, it will address the unmet clinical need to identify biomarkers for early detection of degenerative diseases. This will be significant because it has the potential for a positive impact on the healthcare system, both in terms of improving patient care and reducing the cost of care.

**Figure 1.**
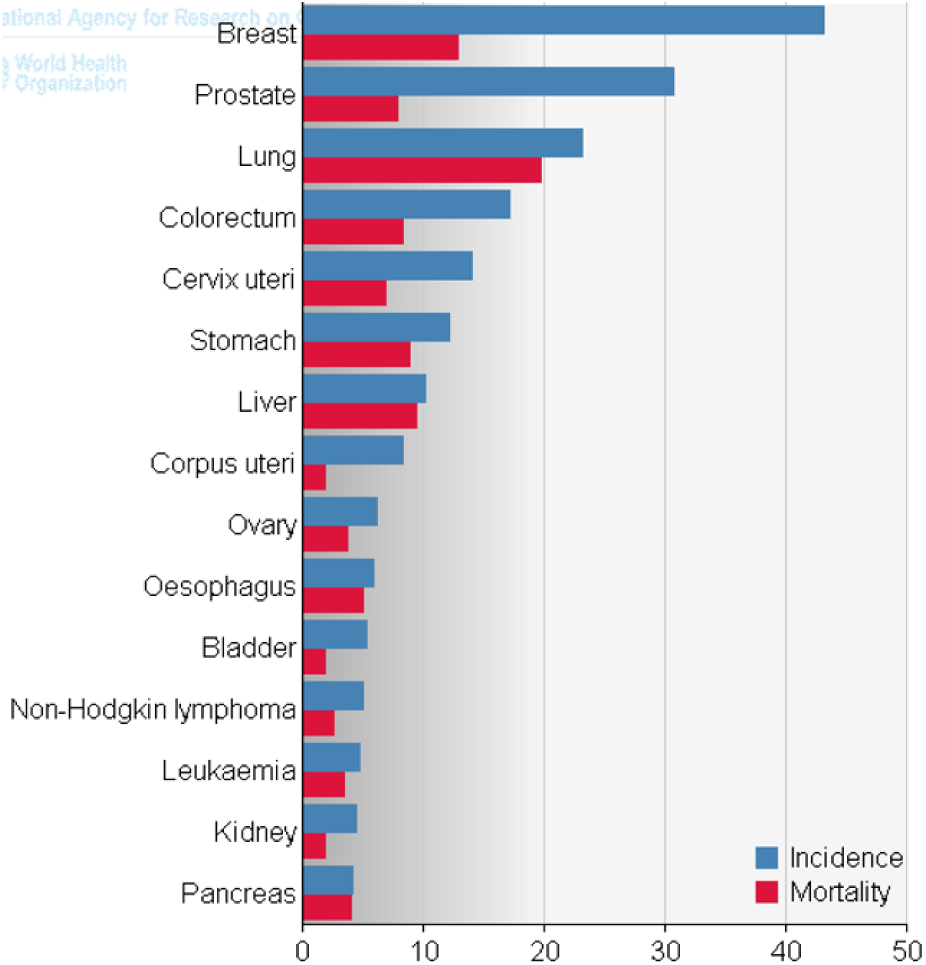
Global annual cancer incidences and mortality rates. Age-standardized rate worldwide per 100,000 inhabitants. The figure demonstrates that for some cancers, such as lung, stomach, liver, esophagus, leukemia and pancreas cancer, the diagnosis is almost inevitably linked to a loss of life. The 5-years survival rate for patients with lung cancer, the “biggest killer” (third from top), is 47% for stage I and as low as 2-20% for stage IV, which underscores the importance of early detection.

In the context of the analysis of samples from individuals diagnosed with a disease and possibly undergoing drug treatments, the significance of the approach will be in providing novel ways for treatment evaluation. Correlation between changes in biomarker levels and treatment responses will allow for early detection of lack of response after treatment and ultimately for optimal drug selection. It will also facilitate the discovery of biomarkers for the prediction of relapse. In the long run, longitudinal analysis of nucleic acids will allow for the development of novel targeted drugs. Recent literature has described many examples of miRs which could be targeted in disease (Duygu et al., 2016; Kim et al., 2017; Singh and Sen, 2017). Therefore, monitoring of their levels in healthy individuals and in patients undergoing disease treatment will likely provide valuable datasets for the pharmaceutical industry for physicians, ultimately allowing them to select the most efficacious treatment sequence and drug combination for each individual patient.

While methods for the early detection of disease based on the analysis of body fluids have become popular, the identification of biomarkers in urine is one of the few completely <u>non-invasive</u> approaches. In addition, it has a clear advantage over other body fluids in the simplicity of collection and processing, as urinary biomarkers are stable for a long period after sample collection. Urine samples with added preservative allow nucleic acid and protein preservation at room temperature for up to a year (see norgenbiotek.com). In addition, panels of miRs in peripheral blood and primary or metastatic tumor samples have been established for several types of tumors and our datasets allowed us to identify many of them in the urine samples we processed (see Appendix). Overall, the analysis of urine samples is a practical way to screen a large number of people at risk of developing a particular degenerative disease, for instance lung cancer.

The objective of the study is to create a biobank that will generate data for research, based on a large number of samples taken, in order to facilitate statistical analyses of systematic changes in our bodies when we become ill. Longitudinal studies of alterations in nucleic acids, such as miRs, will allow for identification of panels of biomarkers of degenerative diseases, such as cardiovascular diseases, cancers and neurological disorders. The biomarker panels will give us the opportunity to perform personalized early detection of disease and alert patients as to the possible onset of a disease before any symptoms are present and prior to any warnings delivered by the current methods developed by the existing healthcare system.

There are approximately two thousand miRs that have been identified in the literature (Kozomara and Griffiths-Jones, 2014) and catalogued in the miRBase database (Kozomara and Griffiths-Jones, 2014). Most of these miRs have been associated with the function of specific human organs and tissues, and the dysregulation of many of them has been linked to cancers (Di Leva et al., 2014; Hayes et al., 2014), cardiovascular diseases (Wojciechowska et al., 2017), neurological disorders (Quinlan et al., 2017) and other degenerative diseases. It has also been demonstrated that it is possible to discriminate between healthy individuals of different ages as well as between the two sexes based on the analysis of miRs isolated in urine samples (Ben-Dov et al., 2016). This work determined that the intra-individual variability is considerably smaller than inter-individual differences, which is important in the context of longitudinal analysis of healthy samples, and that the exact timing of urine collection minimally affects the urine miR transcriptome. Therefore, our <u>hypothesis</u> is that longitudinal changes in miR transcriptomes can be used to detect disease onset before manifestation of any symptoms. As a concrete preliminary analysis of disease-specific changes, we have chosen to highlight the specific case of lung cancer.

Our expectation is that this health monitoring service will alert the participants of the emergence of a particular disease, create the impetus to discuss this alert with her/his physician, and ultimately take appropriate prophylactic action, e.g., to make lifestyle changes or to elect to undergo early treatment. Our <u>central hypothesis</u> is that urinary biomarker panels could serve as personalized health monitoring readouts by virtue of periodic measurements. Furthermore, we hypothesize that gradual changes in urinary nucleic acids will indicate the onset of disease by changes compared to a baseline healthy state that could reveal the type of disease by exhibiting changes in specific biomarkers for each disease. Ultimately, this approach will allow for overall health monitoring and early detection of the onset of any disease, and might also allow for screening for other deviant processes such as physiological disorders.

## MATERIALS AND METHODS

### Study design overview

Our primary methodology is to collect body fluids for analysis. To perform research and identify biomarkers for early detection of disease, every three months we will collect body fluids and store them at negative temperatures with the objective to perform genetic sequencing and statistical analyses of longitudinal data. The urine voids will be collected via specialized urine container 250 ml cups custom-manufactured by Norgen Biotek Corp., which will be shipped via a ready-to-mail enclosed envelope or deposited at a local urine collection point.

### Primary and secondary objectives

Our primary objective is the development of methodology for longitudinal analysis of large cohorts of NGS data based on miRs in body fluids for early detection of disease. This objective will consist of developing a computational analysis pipeline designed specifically to identify meaningful patterns and predictive trends in miR levels present in patient urine specimens. Specifically, we will extend the utility of the algorithms used to generate the preliminary analyses, such as principal component analysis, to integrate consecutive analyses consisting of incremental number of time points and participants in the cohorts. We will particularly aim at detection of disease prior to symptom manifestation to allow for better treatment options by the discovery, in body fluids such as urine, of biomarker panels, which can be used for the early detection of many diseases associated with aging. Our second objective is to discover personalized biomarker panels and allow patients to access pre-clinical research data and make informed decisions about the need for lifestyle changes and/or seeking medical advice. Using information theory, each patient’s data will be analyzed on an individual basis, which will take into account the particularities of each person, and can be parsed in an organ/tissue or disease-specific context. Our aim with this objective will be to provide every individual with the option to make data-driven, informed decisions to prevent the progression of age-related degenerative diseases while slowing down gradual loss of organ function in normal physiology and more acute loss of function in disease.

### Endpoints, inclusion criteria, and sample size

The endpoint of the study is the diagnosis of a degenerative disease (Fig. 2), which provides clinical information and allows us to benchmark our computational predictions. In this context, we present results on the detection of lung cancer in urine samples of 13 stage IV drug-naïve patients, which we compared to 15 healthy individuals without co-morbidities and diseases that affect urinary miRs, such as diabetes and kidney disease. This research study will not have any exclusion criteria. The healthy individuals’ sample collection will be limited between the ages of 45 and 75, while the body fluids collection for individuals already diagnosed with a disease will be universal without any limitations. During the initial data-aggregation phase, when we will rely on a medical diagnosis to establish an endpoint, each participant will be expected to provide information from their general practitioner in regard to the type of diagnosed disease. We anticipate this initial stage to last about two to three years, during which 7% of the participants of age 45 to 75 are expected to develop a degenerative disease. During this stage our focus will be on identifying biomarkers for early disease detection. If we assume ten thousand participants, based on statistics obtained from hospitals (personal communication), within two years 695 participants will develop degenerative diseases of which roughly 220 participants will be diagnosed with a cancer, 436 participants will be diagnosed with a cardiovascular disease and 39 participants will develop a neurological disorder.

**Figure 2.**
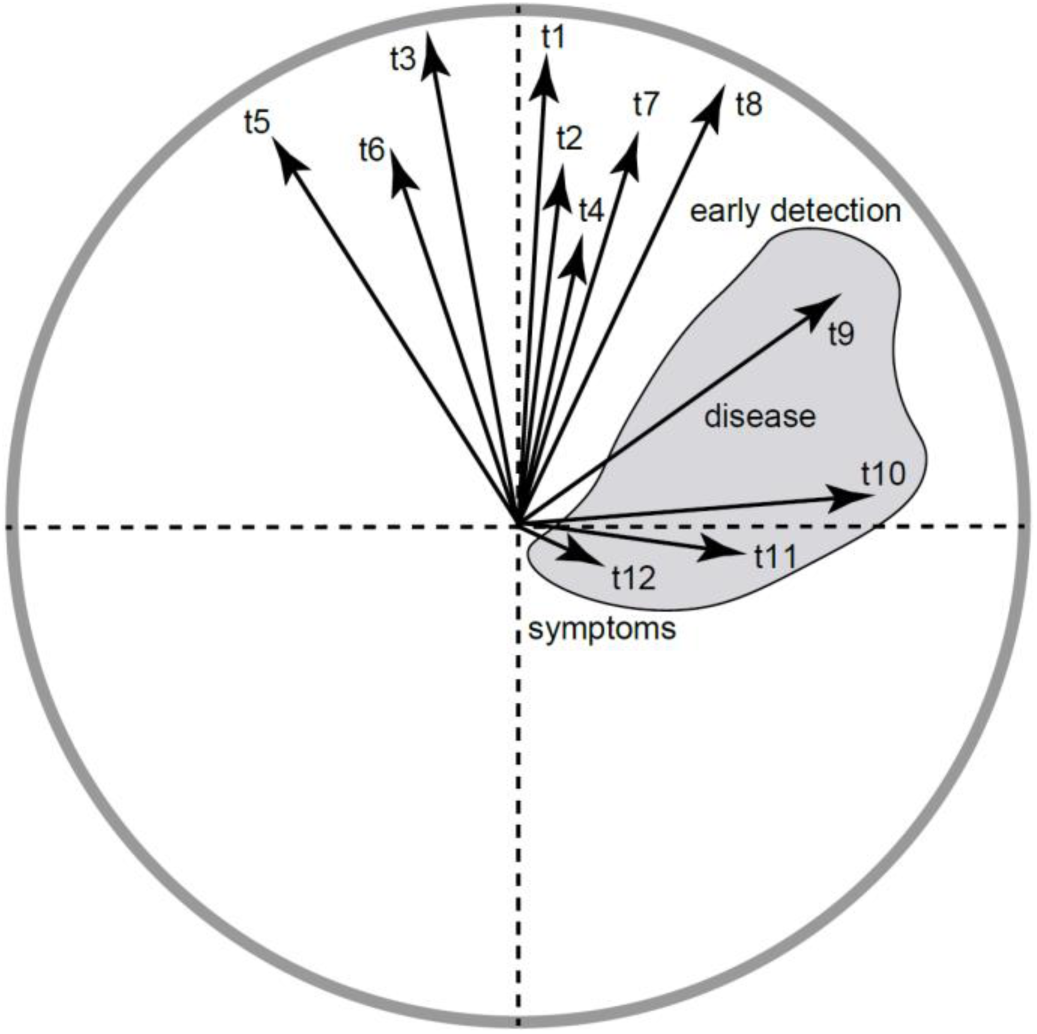
Conceptual figure of early detection of disease using longitudinal data; x-axis PCA1, y-axis PCA2. The numbers represent PCA axes for 12 different time points acquired over three years by taking a sample every three months. Here the first eight samples of the same healthy individual cluster together while for samples taken at time points 9 to 12 the vectors drift further and further away indicating significant changes in the body in disease. Time-point 12 is when the disease is manifested with symptoms, while time-point 9 represents “early detection”.

### Body fluids biobank

To begin to build a repository of patient samples, we will ask participants to contribute body fluids samples (which will be stored in a biobank) and to complete health-related questionnaires every three months. This specific sampling interval is selected because it corresponds to the longest time interval during which new disease can be expected to remain at surgically resectable levels (Quint, 2003). 250 ml of urine void will be collected in a tube containing Norgen Biotek Corp. preservative. Time and date of collection will be logged on a urine worksheet by the laboratory, and the sheet will remain in the tube. Urine appearance will be noted in the log file as “Clear”, “Turbid”, “Yellow” or “Brown”. Before long term storage, the 250 ml collection tube will be centrifuged for 20 min at 4 C at 500 g and the supernatant aliquoted in five 50 ml tubes (the sediment pellet containing epithelial cells will be discarded). For each participant, during each collection, 250 ml of urine void will be aliquoted in five 50 ml Urine Collection and Preservation Tubes (50 cc) and mixed well with the dried preservative for long term storage at -20 C.

### Sample processing

Small RNA were extracted from de-identified urine samples by Norgen Biotek Corp. Our primary methodology is to collect body fluids for analysis. To perform research and identify biomarkers for early detection of disease, every three months we collect body fluids and store them at negative temperatures with the objective to perform genetic sequencing and statistical analyses of longitudinal data. The urine voids are collected via specialized urine container 250 ml cups custom-manufactured by Norgen Biotek Corp. which are shipped at room temperature.

The longitudinal datasets with a disease diagnosis endpoint will allow for the discovery of biomarkers for early detection. Furthermore, the continued collection of samples from these participants will allow for accumulation of datasets for the evaluation of drug efficacy and provide prognostic and predictive value. In the long run, this will empower those participants who develop disease by providing additional information to their treating physicians. The remaining participants (93%) providing healthy samples will continue participation beyond the initial phase of two years and, once early disease biomarkers are discovered and validated, will be offered the chance to participate in our phase two health monitoring service.

### Data analysis

All data analysis programs and graphical representation of the results were developed in R and Matlab. The computer code is available for download at: https://github.com/amatov/DiseaseDetectionUrine. To obtain preliminary data, we used the computational topology software (Perseus (Tyanova and Cox, 2018)) to analyze longitudinal data of 15 healthy donors taken every two months for three different time points. Principal component analysis (PCA) grouped data points of the same individuals together in triplets, which was an expected result, but we were also able to detect trends in how the values for every individual change with time (see Results). This illustrates the transient nature of the miR levels and the importance of understanding normal biological noise / fluctuations in miR levels across the lifespan of healthy individuals.

We also aimed at discovering population-based panels of biomarkers. The small sizes of our preliminary datasets preclude utilization of standard methods and calculate statistical significance, e.g., differential expression analysis (Robinson et al., 2010), so we opted to perform information theory-based computation to identify biomarkers. As the size of our datasets increases, so will our ability to make meaningful biomarker identifications. For the selection of disease-specific panels, several approaches are available for feature selection, for instance, by maximization of mutual information (Jiao et al., 2015) or applying the “maximum of the minimum criterion” (Bennasar M., 2015). The preliminary methodology we utilized for computation of relative entropy via Kullback-Leibler (KL) divergence is based on an adaptive minimax rate-optimal estimator (Han et al., 2016) of the changes in disease from healthy state(s) to cancerous lesion(s) and malignant tumor(s). Consider the KL divergence as:

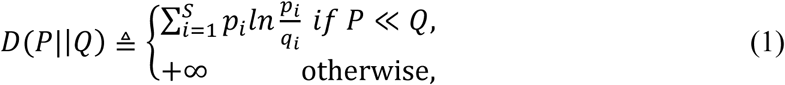

where two patient cohorts are considered, *P* = {*p*_1_, …, *p_s_*} and *Q* = {*q*_1_, …, *q_s_*}, over a common set of miRs of length S (S=947 miRs for this dataset). Testing this approach for significant feature selections on a small cohort of 13 stage IV lung cancer patients’ urine samples, we selected miRs by computing Eq. (1) to identify features for which the tumors differ from healthy samples the most.

All programs for hierarchical clustering analysis and graphical/image representation of microRNAs were developed in R: https://github.com/amatov/DiseaseDetectionUrine. The Kullback-Leibler divergence (Kullback and Leibler, 1951) method used for biomarker selection is described and validated in (Han et al., 2016). The computer code is in Python and is available for download at: https://github.com/amatov/ FragmentomicsSubclinicalDisease.

### Cell culture

Primary and retroperitoneal lymph node metastatic prostate tumor and sternum metastatic rectal tumor tissues were dissociated to single cells using modified protocols from the Witte lab. Organoids were seeded as single cells in three 30 µL Matrigel drops in 6-well plates. Organoid medium was prepared according to modified protocols from the Clevers lab and the Chen lab. We expressed fluorescently tagged MT markers in the organoids by using lentivirus-mediated transduction and had best results infecting organoids in an exponential growth phase. To optimize imaging conditions, blasticidin selection was tuned such that only 20-30% of organoid cells are GFP-labeled (Matov, 2025). This way, we achieved better contrast of imaging end-binding proteins 1 (EB1) dynamics.

To obtain a single cell suspension, tissues were mechanically disrupted and digested with 5 mg/ml collagenase in advanced DMEM/F12 tissue culture medium for several hours (between 2 and 12 hours, depending on the biopsy/resection performed). If this step yielded too much contamination with non-epithelial cells, for instance during processing of primary prostate tumors, the protocol incorporated additional washes and red blood cell lysis (Goldstein et al., 2011). Single cells were then counted using a hemocytometer to estimate the number of tumor cells in the sample, and seeded in growth factor-reduced Matrigel drops overlaid with prostate cancer medium (Gao et al., 2014). With radical prostatectomy specimens, we had good success with seeding three thousand cells per ∼30 μl Matrigel drop, but for metastatic samples organoid seeding could reliably be accomplished with significantly less cells, in the hundreds. To derive organoids from patient circulating tumor cells (CTCs), liquid biopsy samples of 40 ml peripheral blood would be collected, processed, and plated in a Matrigel-Collagen-Fibronectin matrix to form organoids similarly to the metastatic breast cancer organoids we cultured from mouse CTCs (Matov, 2024a).

To transduce organoids, we modified protocols from the Clevers lab to adapt to the specifics of prostate organoid culture (such as the significant differences in proliferation rates in comparison to colon and rectal organoids). We found out empirically that cells in mid-size organoids (60-100 µm in diameter) infect at much better rates than trypsinized single organoid cells. These were the steps we followed to express EB1ΔC-2xEGFP in organoids: (1) Add Dispase (1 mg/ml) to each well to dissolve the Matrigel at room temperature for 1 hour. (2) Spin down (at 1,000 rpm for 4 minutes) and mix organoids with 10 µl of viral particles (enough for 1 well with three Matrigel drops of 30-40 µl with organoids containing 1-2 million cells) with Y27632 ROCK inhibitor and Polybrene (1:1,000) for 30 minutes. (3) Spin the organoids with viral particles for 1 hour at 600g. (4) Leave the organoids for a 6-hour incubation. (5) Spin down and plate in Matrigel. (6) 1 hour later, add medium. We used blasticidin (1:20,000) for only one round of medium (3 days), because an increase of the density of labeled cells in the organoids reduced our ability to image microtubule ends with good contrast.

### Microscopy imaging

Organoids were imaged using transmitted light microscopy at 4x and 20x magnification, and phase contrast microscopy at 20x magnification on a Nikon Eclipse Ti system with camera Photometrics CoolSnap HQ2.

## RESULTS

### Nucleic acids biomarkers

At least 1,917 human miRs have been described in the literature (Kozomara and Griffiths-Jones, 2014), and we have detected 947 of them in urine. The levels of miRs change as a function of age and systematic differences exist between the two sexes (Ben-Dov et al., 2016). There are three main categories of degenerative diseases - cardiovascular (e.g., hypertension, coronary disease, and myocardial infarction), neoplastic (tumors and cancers), and nervous system-related (e.g., Alzheimer’s disease and Parkinson’s disease). The etiology of these diseases typically involves some combination of aging and poor lifestyle choices. It is, therefore, conceivable that the collection and analysis of urinary miR panels longitudinally would allow to delineate changes in physiology associated with a particular disease in its early (asymptomatic) stage.

Figure 3 shows an example of the miRs detection in the urine of a healthy individual. Not all of the miRs we detected in urine are present in all subjects for each of the time points. We aimed at discovering personalized biomarker panels, which would allow anyone to make informed decisions and alert about the need for lifestyle changes or to seek medical advice. In this context, we sought to identify meaningful patterns and predictive trends in miR levels present in patient urine and blood specimens.

**Figure 3.**
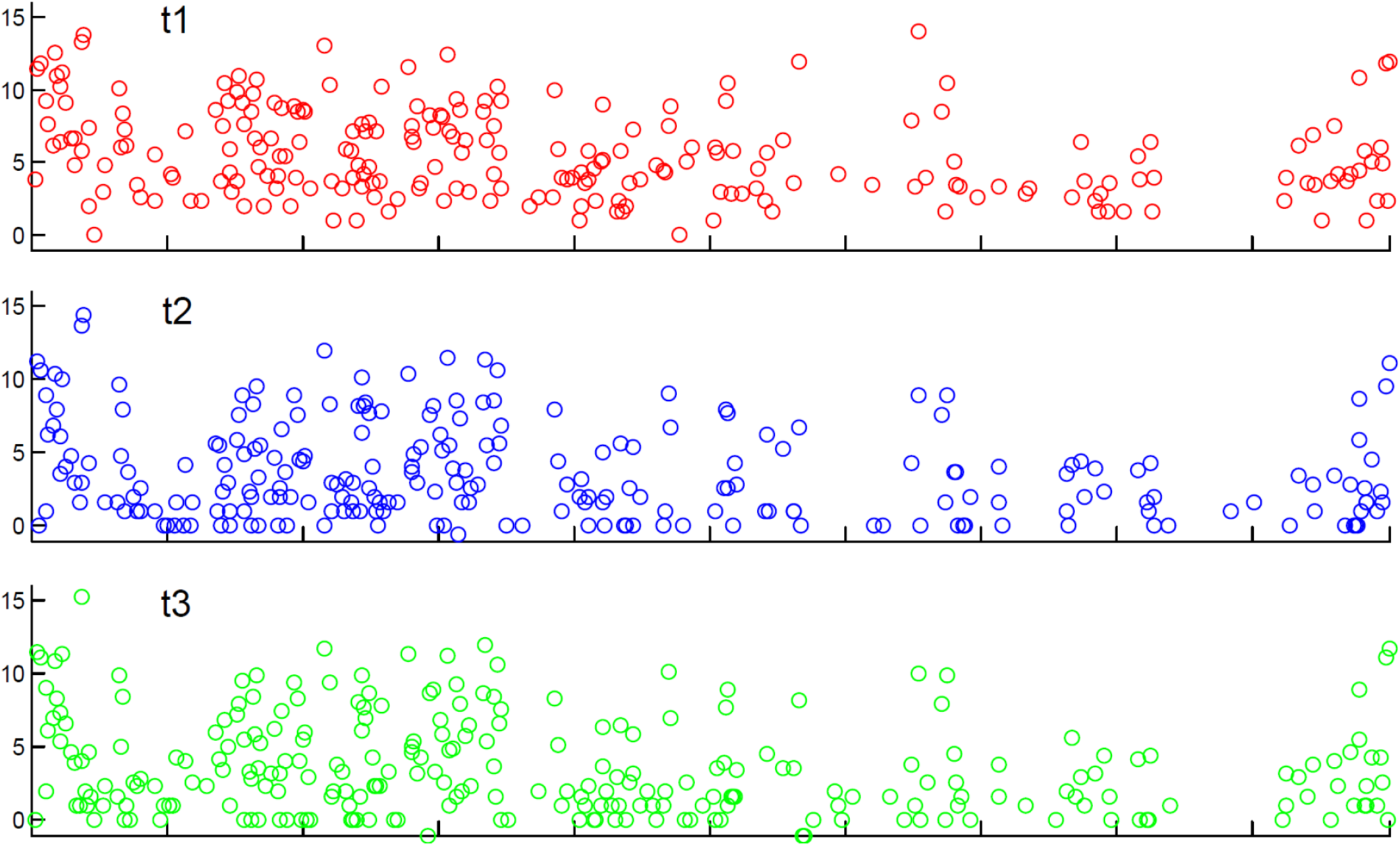
Expression levels (logarithmic scale) of 947 miRs sequenced by NGS from urine samples for one of the 15 healthy individuals over three time points. Example of visualization of urine data based on 947 miRs plotted on the X-axis versus miR expression levels (in arbitrary units) as plotted on the Y-axis. Time point 1 levels are displayed in red, time point 2 levels are displayed in blue and time point 3 levels are displayed in green.

### Biomarker panels in the literature

The treatment of lung cancer is often hampered by late diagnosis (Del Ciello et al., 2017; Ellis and Vandermeer, 2011; Polanski et al., 2016). When discovered early, the disease is curable via surgical intervention as adenocarcinoma nodules can simply be resected (Lackey and Donington, 2013). For patients with locally advanced or metastatic disease, multiple drug treatment options are offered yet there are rarely curative regimens available (Socinski et al., 2018). Immunotherapy seems to be a promising approach for a curative therapy (Rizvi et al., 2015), but it too has its limitations as there are very few patients who respond to it and the associated toxicity is of considerable concern (Carbone et al., 2017). While current research efforts in the area of drug development have focused on identifying biomarkers for immunotherapy susceptibility (Ayers et al., 2017; Cristescu et al., 2018; Gettinger et al., 2018), our approach to improving the standard of care in lung cancer treatment is to identify biomarkers for early detection of the disease when curative surgery is still a viable option.

Approximately 190 miRs (Aushev et al., 2013; Backes et al., 2016; Bianchi et al., 2011; Boeri et al., 2012; Boeri et al., 2011; Cazzoli et al., 2013; Chen et al., 2012b; Geng et al., 2014; Leidinger et al., 2014; Leidinger et al., 2016; Li et al., 2015; Markou et al., 2016; Nadal et al., 2015; Nadal et al., 2014; Qu et al., 2017; Wang et al., 2014; Wozniak et al., 2015; Yan et al., 2014; Zhou et al., 2017; Zhu et al., 2016) have been linked to lung cancer based on transcriptomics data of specimens derived from primary tumors and/or blood. The top 25 most common markers have been identified in at least three independent screens and some are listed in as many as six or more, suggesting that they are disease markers of high fidelity. The fidelity of markers will also be based on whether they have been identified in both the primary tumors and in blood samples and we will start the health monitoring of lung cancer by accumulating statistics on these 190 markers (see Appendix) with a special emphasis on the top 25 most established ones.

In addition to establishing this miR biomarker panel for lung cancer, we have also identified similar but smaller (i.e., based on fewer established blood biomarkers) panels (see Appendix) for colorectal and prostate cancer (in males); breast, uterus, cervical, and colorectal cancer (in females); as wells as urinary pancreatic cancer biomarkers (Debernardi et al., 2015). The necessity to consider different sets of biomarkers for males and females, depending on age will be discussed in the next section. The biomarker lists will be updated as new literature becomes available and whenever our own work allows for the identification of novel markers.

### Published lung cancer miR biomarkers found also in urine

Lung cancer is the leading cause of cancer-related deaths and claims more lives each year than all other major cancers combined and lung cancers are generally diagnosed at an advanced stage because patients lack symptoms in the early stages of the disease. Based on our literature search, we identified a panel of 25 high fidelity lung cancer biomarkers, based on miRs each previously linked to lung cancer development, progression and drug resistance in multiple papers (between three and eight; see Appendix) on the analyses of blood of lung cancer patients and primary lung tumors (Aushev et al., 2013; Backes et al., 2016; Bianchi et al., 2011; Boeri et al., 2012; Boeri et al., 2011; Cazzoli et al., 2013; Chen et al., 2012b; Geng et al., 2014; Leidinger et al., 2014; Leidinger et al., 2016; Li et al., 2015; Markou et al., 2016; Nadal et al., 2015; Nadal et al., 2014; Qu et al., 2017; Wang et al., 2014; Wozniak et al., 2015; Yan et al., 2014; Zhou et al., 2017; Zhu et al., 2016). Below, we are providing information on the effects on lung physiology of these 25 biomarkers.

We measured an increase in the levels of 16 of these 25 miRs for one of the healthy individuals in our baseline cohort of 15 people (Fig. 4). We did not consider those miRs for which there have been reports in the literature of a downregulation in disease, because their levels might appear decreased in urine for other physiological reasons than cancers. The most commonly published biomarkers of lung cancer, which we found to show an increase longitudinally are miR-21-3p, miR-140-3p and miR-93-3p. Nicotine-induced miR-21-3p promotes chemoresistance in lung cancer by negatively regulating FOXO3a (Zhang et al., 2022b). miR-140-3p is the most stably expressed plasma exosomes miR in lung cancer patients (Jiang et al., 2024). miR-93 is up-regulated in a wide variety of malignancies, such as lung cancer and can serve as a biomarker for drug resistance (Hussen et al., 2023). This indicates that the longitudinal analysis of urinary miRs in healthy individuals has the potential for providing biomarkers for early disease detection.

**Figure 4.**
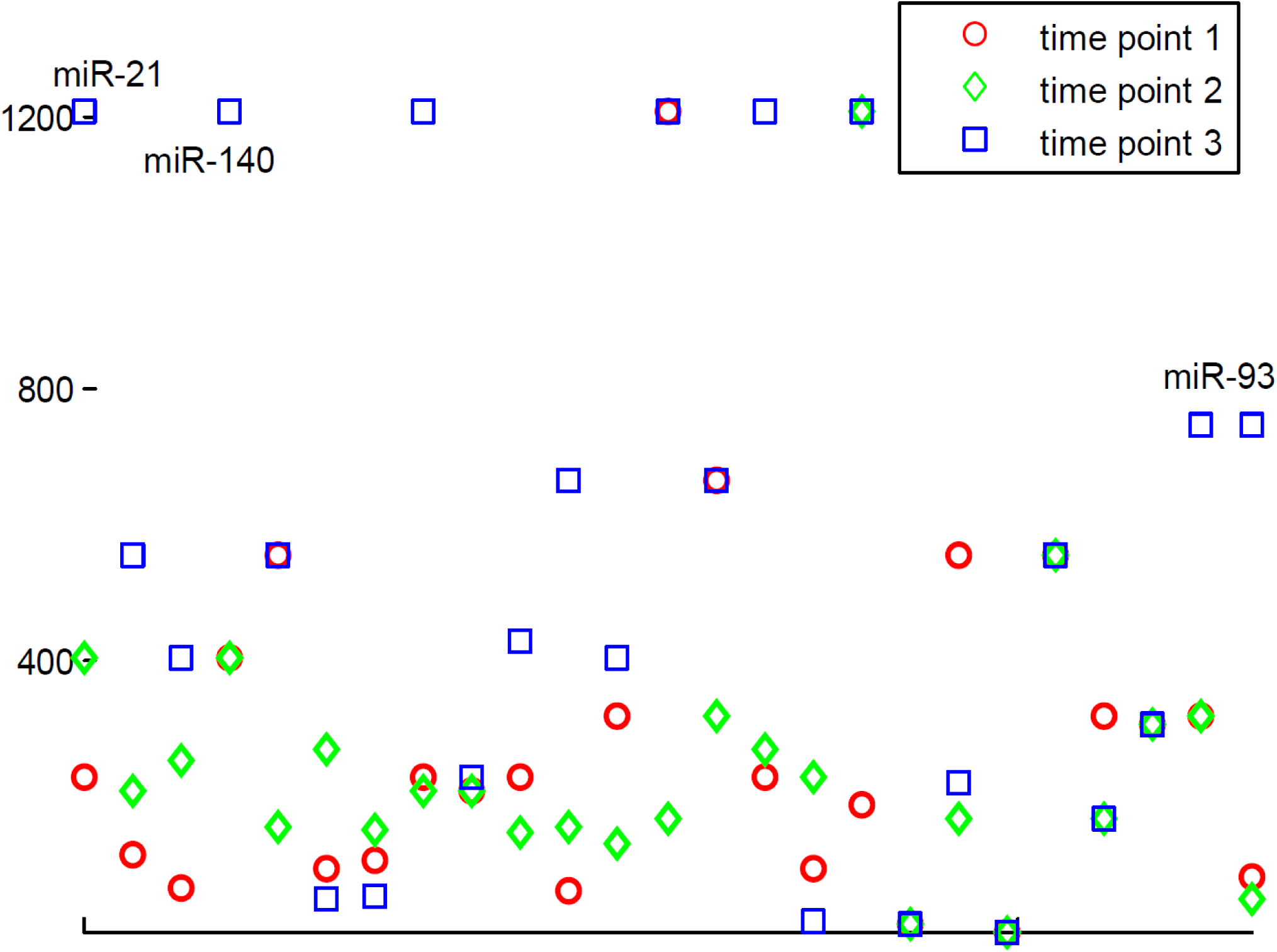
Longitudinal changes of lung cancer-related urinary miRs in a healthy individual. An example of up-regulation of 16 urinary biomarkers within a panel of 25 high fidelity lung cancer biomarkers, based on miRs each previously linked to lung cancer development, progression and drug resistance in multiple (between three and eight) papers on the analyses of blood of lung cancer patients and primary lung tumors. X-axis, list of the biomarkers: from left to right: (1) miR-21-3p, (2) miR-21-5p, (3) miR-140-3p, (4) miR-140-5p, (5) miR-155, (6) miR-200b-3p, (7) miR-200b-5p, (8) miR-223-3p, (9) miR-223-5p, (10) miR-221-3p, (11) miR-221-5p, (12) miR-145-3p, (13) miR-145-5p, (14) miR-150-3p, (15) miR-150-5p, (16) miR-200a-3p, (17) miR-200a-5p, (18) miR-205-3p, (19) miR-205-5p, (20) miR-210-3p, (21) miR-210-5p, (22) miR-339-3p, (23) miR-339-5p, (24) miR-93-3p, (25) miR-93-5p. Y-axis, abundance levels; we performed data normalization using the quantile normalization method recommended for next-generation sequencing data based on single color experiments.

### Computational analysis

#### Principal component analysis of healthy longitudinal samples

After establishing that it was feasible to identify published cancer miRs in urine, we sought to investigate whether healthy donors retain similar patterns in their miR profiles longitudinally via PCA (Fig. 5). To this end, we processed samples of 15 healthy individuals collected every two months for three time points, i.e., 45 samples in total. This analysis showed that even if there is a certain level of variability, (i) each three longitudinal samples from the same healthy individual cluster together - at least for two of the three time points - and (ii) the longitudinal sample triplets for each of the different healthy individuals form separate clusters (see on Fig. 5 sample triplets per individual color-coded in the same color – pink for donor #1, red for donor #2, cyan for donor #3, blue for donor #4, brown for donor #5, yellow for donor #6, orange for donor #7, etc.). This demonstrates that, in normal physiology, urinary miR panels can identify the same person longitudinally.

**Figure 5.**
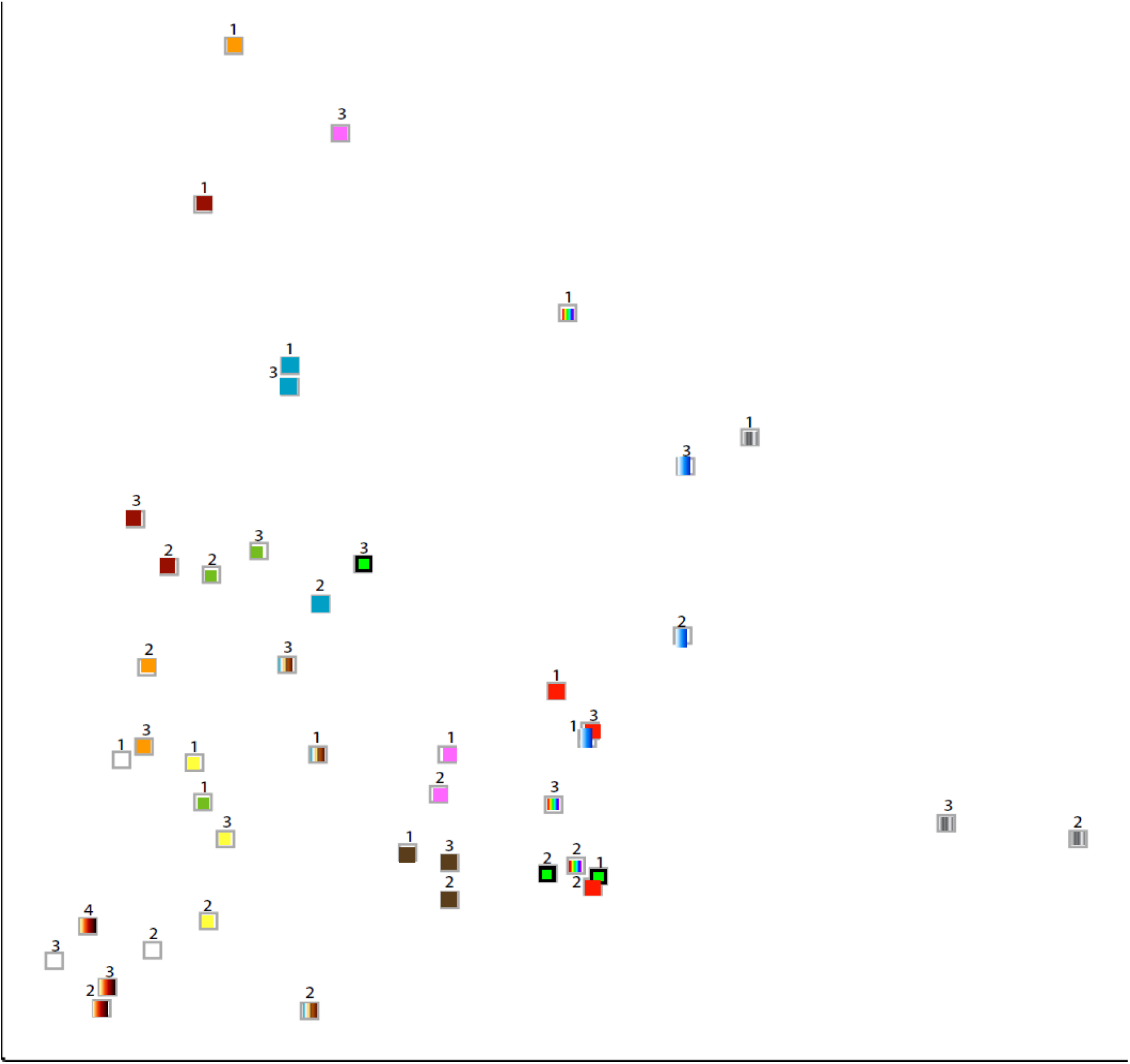
Longitudinal analysis of 45 samples from 15 healthy donors; X-axis Component 1 (21.4%), Y-axis Component 2 (13.1%). PCA analysis of longitudinal data of healthy donors taken at three different time points demonstrates the transient nature of the miRs levels. The panel consists of 15 healthy donors which provided void urine samples in the Netherlands. Each triplet 1-2-3 in the same color on the PCA scatter plot belongs to the same healthy individual over three time points. The time step at which these samples were collected and sequenced was two months. One of the triplets is labeled 2-3-4 because the first sample was not processed correctly and that required the collection of another one at a later, fourth, time point.

#### Principal component analysis of lung cancer stage IV drug-naive samples

We used a PCA library FactoMineR (Le S., 2008) miRs also allowed to perform multivariate analysis (Fig. 6) for stratification of patient cohorts based on different characteristics such as age, sex, and smoking status individually or in any combination. The separation between the groups is expressed in the angular orientation of the vectors as well as in the length (amplitude). The labeling is as follows: patient (p) or control/health (c), next is the sample number, next is the gender (m/f) followed by age and smoking status – nonsmoker (n) or not provided (x), which we assume to represent the smokers. On the lower left quadrant we see a clear cluster (Cluster I) of three assumed smokers of age 70-80 years. Up, vertical there are two male smokers in their mid-50s (Cluster II). In the upper left quadrant there are four (three of them very short) vectors representing a subset of participants of age mid-60s until 70 (Cluster III). There are three vectors pointing to the right of people (possibly all nonsmokers) of age 71 and 72 years (Cluster IV), which can also be associated with c9m71n (same age). In the lower right quadrant, there is a cluster of three healthy people, all nonsmokers, of age 48-50 years (Cluster V).

**Figure 6.**
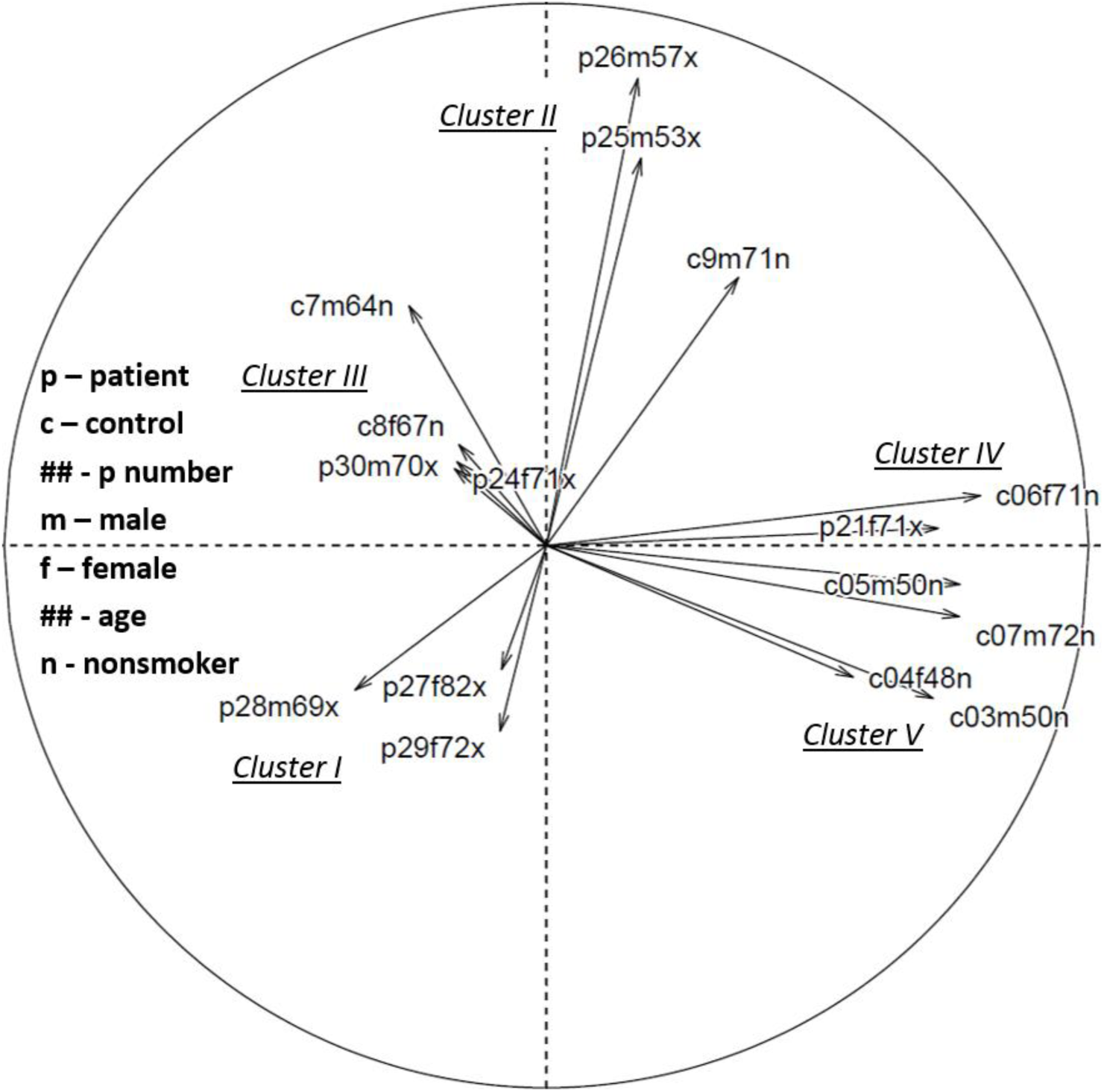
Multivariate clustering of lung cancer patients and healthy people urine data; x-axis PCA1, y-axis PCA2. The separation between the groups is expressed in the angular orientation of the vectors as well as in their length (amplitude). The labeling is as follows: patient (p) or control/healthy (c), next is the two-digit sample number, next is the gender (m/f) followed the age and smoking status – nonsmoker (n) or not provided (x), which we assume to represent the smokers.

#### Identifying disease biomarkers by Kullback–Leibler divergence

One of our objectives has been to discover personalized disease-specific miRs biomarker panels based on significant changes in organ or tissue regulation in disease. As a first step in this regard, we aimed at discovering population-based panels of biomarkers. The preliminary methodology we utilized for computation of relative entropy via Kullback-Leibler (KL) divergence is based on an adaptive minimax rate-optimal estimator (Han et al., 2016) of the changes in disease from healthy state(s) to cancerous lesion(s) and malignant tumor(s). We set our threshold at 1.2 bits divergence empirically and this allowed us to identify 20 biomarkers discriminative of lung cancer (Fig. 7). All 20 biomarkers have previously been published in the literature on lung cancer based on analyses of primary lung tumors and blood from lung cancer patients (Bao et al., 2018; Wan and Zheng, 2021; Wang et al., 2014). This selection demonstrates the suitability and ability of this method for the identification and selection of disease-specific biomarkers. Even if this computation is based on small sample cohorts and it is not patient-specific, it indicates, for the first time, the possibility to detect lung cancer in urine samples. Our objective will be to provide every patient with the option to make data-driven decisions in the context of the prevention of the progression of lung cancer and other degenerative diseases.

**Figure 7.**
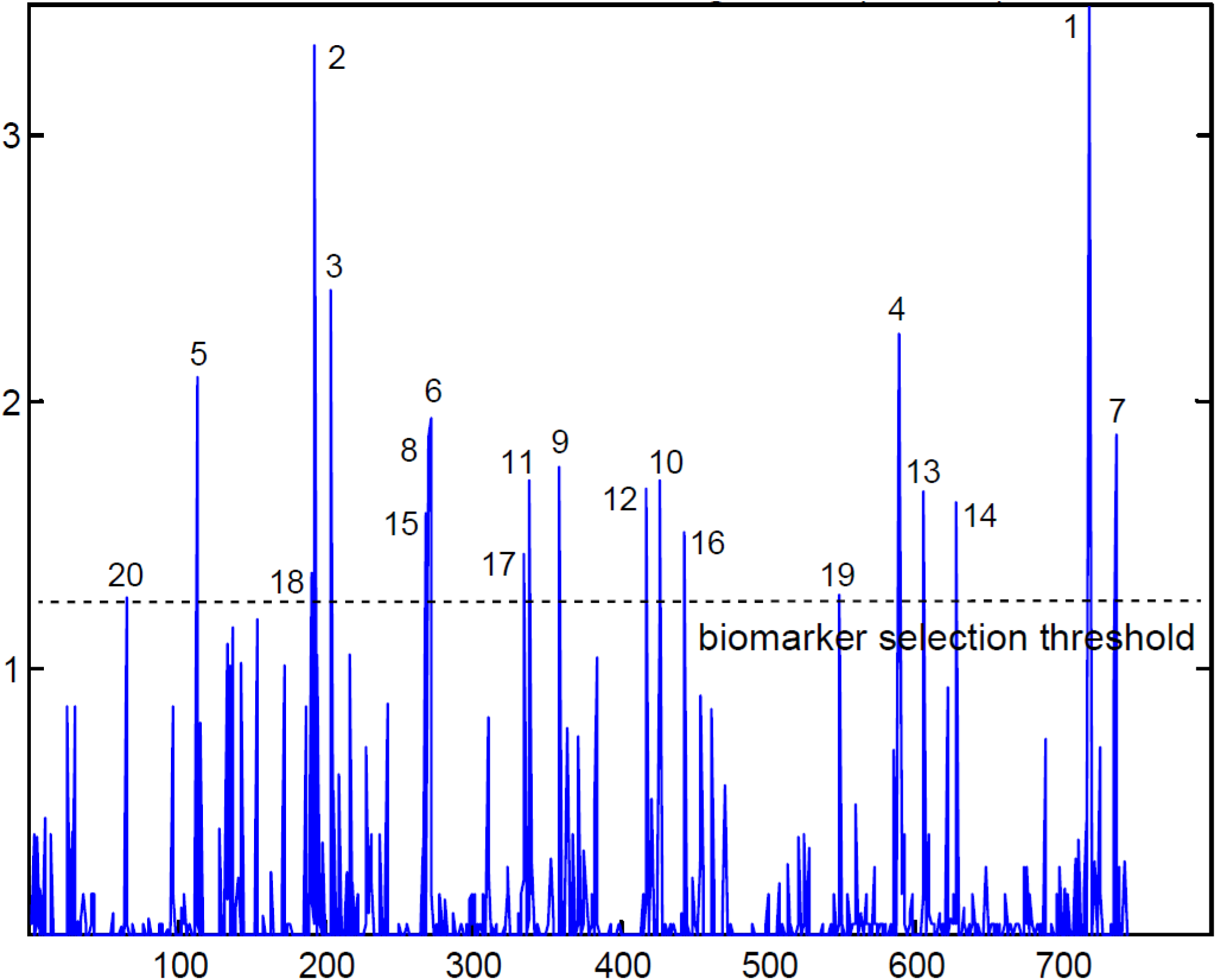
Kullback-Leibler divergence between drug-naïve lung cancer patients and healthy individuals. miRs levels in a cohort of 28 participants (13 stage IV drug-naïve lung cancer patients and 15 healthy individuals) allowed for the selection of a panel of 20 discriminative biomarkers out of 947 (743 in these two cohorts) detected by next-generation sequencing in urine samples: (1) miR-891a-5p, (2) miR-196a-5p, (3) miR-200a-5p, (4) miR-577, (5) miR-141-3p, (6) miR-29c-3p, (7) miR-95-3p, (8) miR-29b-3p, (9) miR-361-5p, (10) miR-429, (11) miR-335-5p, (12) miR-421, (13) miR-628-3p, (14) miR-660-5p, (15) miR-29a-3p, (16) miR-4454, (17) miR-330-3p, (18) miR-194-5p, (19) miR-532-3p, (20) miR-1271-5p. All 20 biomarkers have previously been published in the literature on lung cancer based on analyses of primary lung tumors and blood from lung cancer patients.

Computing KL divergence has been proposed as an approach for detecting sudden deterioration of complex diseases by dynamical network biomarkers (Chen et al., 2012a), which can be accomplished with the use of one patient sample. The distribution-embedding scheme transforms the data from the observed state-variables with high level of noise to their distribution-variables with low level of noise, and thus significantly reduces signal fluctuations (Liu et al., 2015). Increasing the dimensions of the observed data by moment expansion that changes the system from state-dynamics to probability distribution-dynamics allows the derivation of new data in a high-dimensional space, but with much lower noise levels. Single-sample KL divergence has thus been proposed as an approach to detect the early-warning signal of critical transition during the progression of lung cancer. The algorithm was applied to datasets of lung squamous cell carcinoma, lung adenocarcinoma, and acute lung injury (Zhong et al., 2020). As the method identifies the critical state (or tipping point) at a single sample level and identifies signaling biomarkers, it can be of great potential in personalized pre-disease diagnosis in lung cancer. The genes it selected were involved in lung cancer signaling and cellular processes, such as cytoskeleton organization, chromosome condensation, regulation of cell division, programmed cell death, among others (Zhong et al., 2020), demonstrating a wide clinical applicability of KL divergence calculations.

#### Function of discriminative biomarkers identified in urine

To establish the clinical significance of the 20 urinary biomarkers we discovered, our next step was to identify their concrete function based on publications on peripheral blood and tumor tissue samples. Below is information on the effects of the biomarkers on tumor physiology; biomarkers are listed based on their divergence levels of healthy individuals from lung cancer patients in the cohorts we analyzed, as shown on Fig. 7. In addition, we demonstrate the discriminative effectiveness of some of the biomarkers, which for miR-200a-5p has a Youden index of 0.85 (Fig. 8).

**Figure 8.**
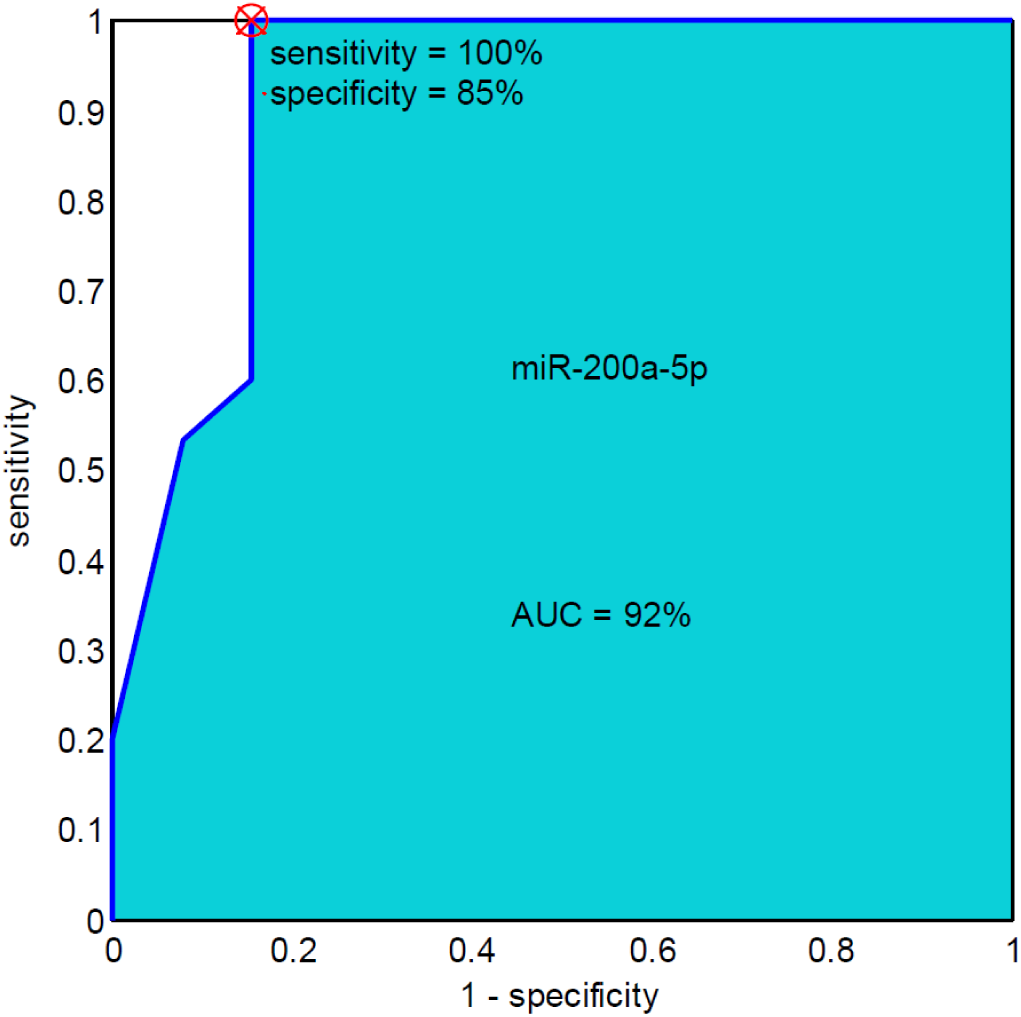
Receiver operating characteristic curve analysis. miR-200a-5p has previously been published in four different papers on the analyses of blood of lung cancer patients and primary lung tumors (see Appendix). Red asterisk indicates the highest Youden’s score index (sensitivity + specificity-1). AUC, area under the curve.

**miR-891a-5p** downregulation results in suppressed tumor cell migration, proliferation and invasion, while the upregulation of miR-891a-5p leads to enhanced tumor cell activity and promotes tumorigenesis non-small cell lung cancer (NSCLC) (Wan and Zheng, 2021).

**miR-196a-5p** is a putative diagnostic biomarker and its expression levels are increased significantly in NSCLC tissues compared with non-tumor adjacent normal tissues (Bao et al., 2018).

**miR-200a-5p** exhibits a tumor suppressive role by regulating proliferation and apoptosis in NSCLC, and could target RHPN2 (Huang et al., 2020).

**miR-577** inhibits cell proliferation and invasion in NSCLC (Men et al., 2019).

**miR-141-3p** promotes the proliferation of NSCLC (Liu et al., 2019) and reduces pulmonary hypoxia / reoxygenation injury (Zhan et al., 2024).

**miR-29c-3p** is significantly decreased in the plasma of lung cancer patients, especially in those with early-stage lung cancer, and can be used as a biomarker discriminating between NSCLC and small cell lung cancer (SCLC) (Zhang et al., 2023).

**miR-95-3p** inhibits the invasiveness of metastatic lung cancer through downregulation of cyclin D1 (Hwang et al., 2015).

**miR-29b-3p** reverses cisplatin resistance by targeting COL1A1 in NSCLC cells (Jia and Wang, 2020).

**miR-361-5p** plays an oncogenic role in lung cancer through the regulation of SMAD2 (Othman and Nagoor, 2019).

**miR-429** promotes the proliferation of NSCLC cells via targeting DLC-1 (Xiao et al., 2016).

**miR-335-5p** is significantly decreased in parenchymal lung fibroblasts of smokers (Ong et al., 2019).

**miR-421** confers paclitaxel resistance by binding to the KEAP1 3’-untranslated region and predicts poor survival in NSCLC (Duan et al., 2019).

**miR-628-3p** promotes apoptosis and inhibits migration in lung cancer cells by negatively regulating *HSP90* (Pan et al., 2018).

**miR-660-5p** plays a role in lung tumorigenesis and could be targeted in lung cancer therapy (Borzi et al., 2017).

**miR-29a-3p** prevented NSCLC tumor growth and cell proliferation, migration, and invasion by inhibiting the Wnt/β-catenin signaling pathway (Zhang et al., 2022a).

**miR-4454** has the potential to be used as a biomarker for early diagnosis of lung cancer (Feng et al., 2023).

**miR-330-3p** promotes metastasis and epithelial-mesenchymal transition via GRIA3 in NSCLC (Wei et al., 2019).

**miR-194-5p** downregulates the expression of RAC1 and can prevent metastasis in NSCLC (Ni et al., 2021).

**miR-532-3p** inhibits metastasis and proliferation of NSCLC by targeting FOXP3 (Ni et al., 2021).

**miR-1271-5p** has the potential to be used as a biomarker for early diagnosis of lung cancer (Zhang et al., 2024).

Several of these 20 miRs have been shown to regulate other organs than the lungs as well. This study, to our knowledge, is the first investigation that indicates the ability to detect a panel of miRs as lung cancer biomarkers in urine. Figure 9 shows the results of PCA of the patient and healthy cohorts based on the 20 biomarkers only. To improve the separation, we have added color-coding of miR-532-3p, which improves the result for patients #4 and #13.

**Figure 9.**
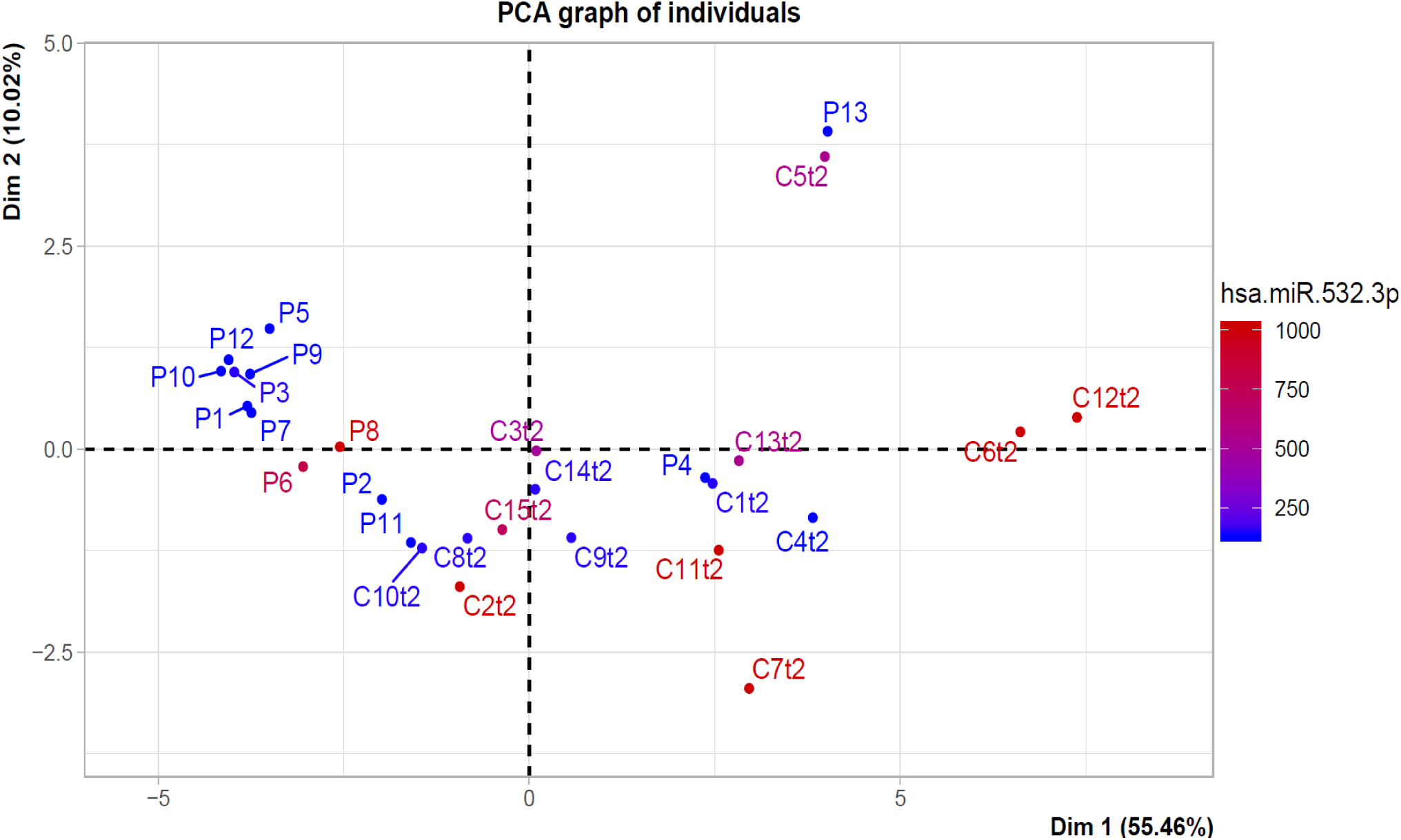
Principal component analysis plot demonstrates spatial separation between patients and healthy individuals. Analysis based on a panel of 20 miRs selected via KL divergence out of 743 miRs sequenced by NGS. An additional separation between lung cancer patients and healthy individuals is provided by color-coding of miR-532-3p levels. P1-P13 denotes the 13 lung cancer patients, C1t2-C15t2 denotes the 15 healthy (control) individuals in time-point 2 (out of 3), i.e., the middle time-point. All patient samples, besides for P4 and P13, clearly cluster to the left side of the plot. In addition, the level of miR-532-3p separates P4 and P13 from most of the control samples.

## DISCUSSION

The cancer incidences and mortality rates worldwide demonstrate that for some cancers, such as lung, stomach, liver, esophagus, leukemia and pancreas cancer, the early diagnosis is of critical importance. We will develop algorithmic solutions to facilitate the longitudinal analyses of transcriptomic miR data from tens of thousands of patients (about 1,917 miRs have been described in the literature and we have detected 947 of them in urine) and the early detection of disease. To achieve this end, we will use multivariate data and multi-dimensional clustering analyses as well as information theory approaches based, for instance, on relative entropy computation.

During treatment of lung cancer, the approach presented in this contribution can be combined with an interrogation of patient-derived lung cells, such as circulating tumor cells (CTCs) (Fig. 10) and organoids (Fig. 11) (Matov, 2024d). To isolate CTCs in peripheral blood, microfluidic techniques (Gleghorn et al., 2010) combine size and surface specificity to exploit differences between CTCs and erythrocytes - but capture many leukocytes (Fig. 10A). Methods for CTC enumeration that do not utilize any enrichment are an alternative for collecting all CTCs within a sample (Fig. 10B). Specialized microfluidic devices for hydrogel-based capture and light-induced release of viable CTCs can lead to high purity and high yield CTC isolation (Applegate et al., 2004; LeValley et al., 2019; Liu et al., 2022).

**Figure 10.**
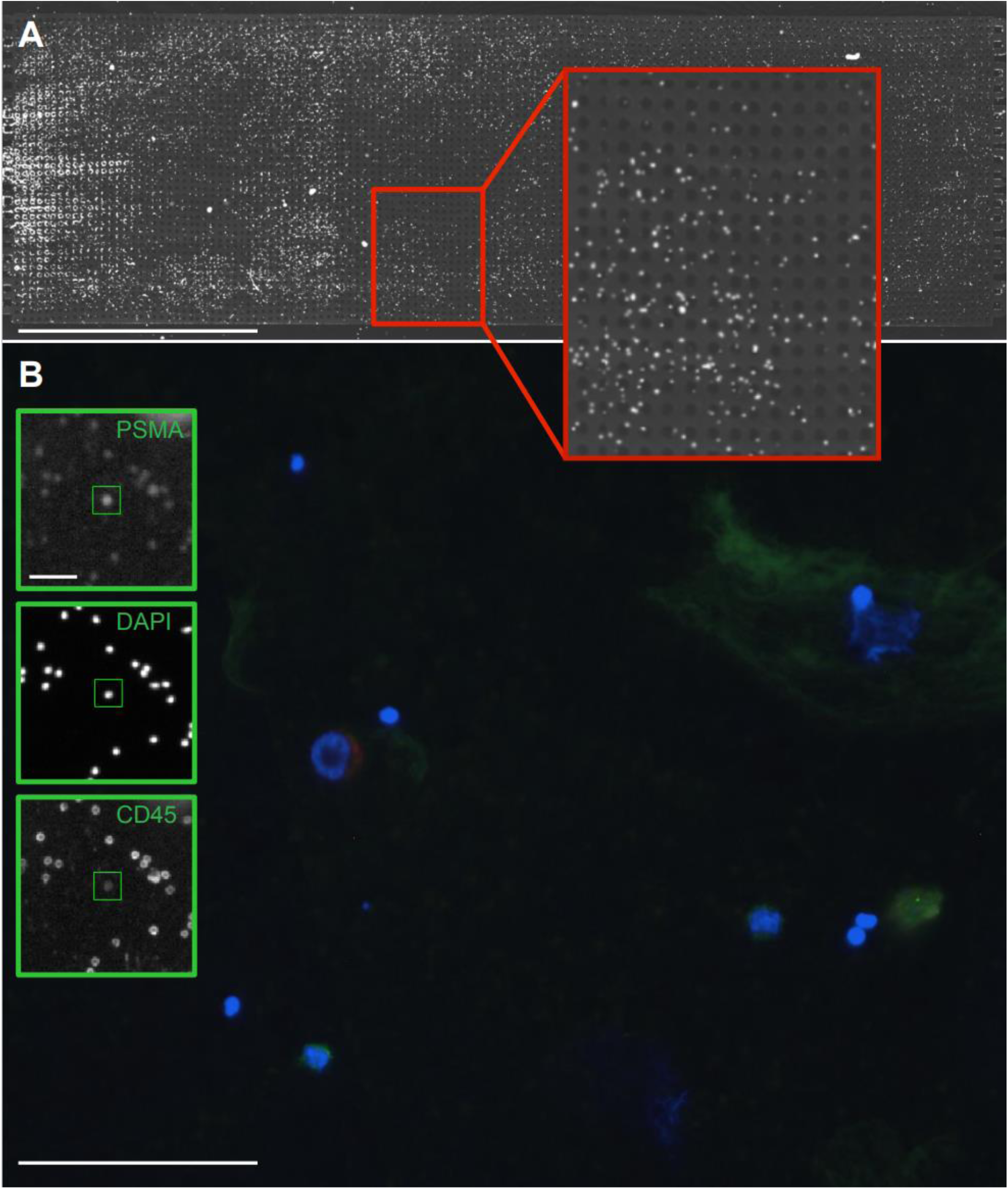
Circulating tumor cells captured with or without an enrichment method. (A) Microfluidic device for capture of live CTCs. Scale bar equals 7 mm. Inset shows zoom-in of a small area of the chip to better see the captured cells, almost all of which are leukocytes. (B) Cells imaged on a coverslip without sample enrichment. Figure legend: PSMA (HuJ591, tumor marker, red), CD45 (leukocyte marker, green), DAPI (nuclear marker, blue). The diameter of the patient CTC in the middle of the image is about 30 µm. The scale bar equals 100 µm. Three insets show the automated detection of a CTC and the three imaging channels: PSMA, DAPI, and CD45 (magnification 4x). Scale bar equals 30 µm.

**Figure 11.**
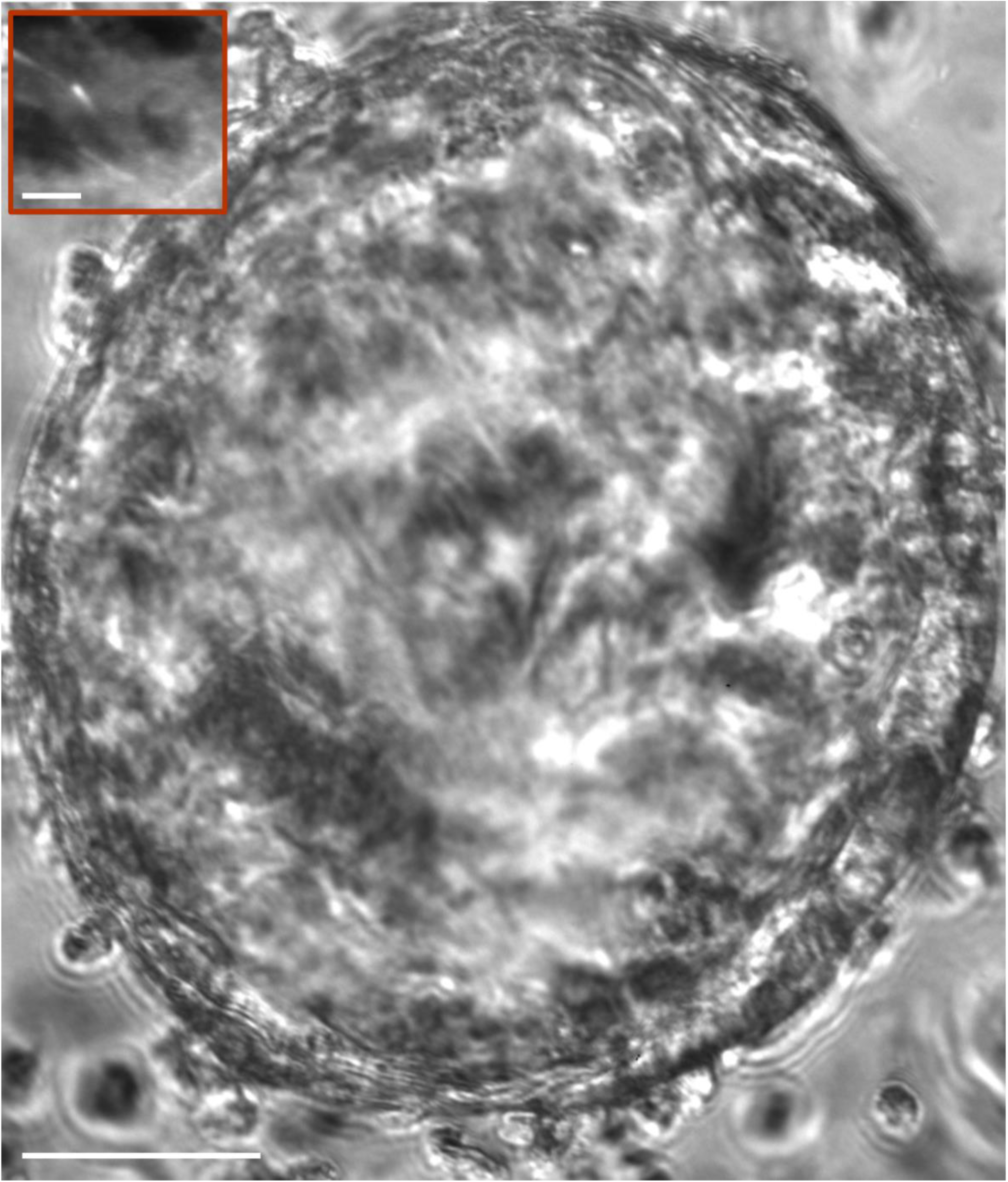
Lung organoid derived from a 70 mg chest-wall resection of a patient with metastatic rectal cancer. Day 25. Magnification 20x, transmitted light microscopy. Scale bar equals 150 µm. The inset shows fluorescent EB1 comets at the tips of polymerizing microtubules. Scale bar equals 1 µm. See also Video 1.

Prostate-specific membrane antigen (PSMA), a cell surface glycoprotein that is commonly overexpressed by prostate cancer cells relative to normal prostate cells, provides a validated target in tumors of the prostate (Bander et al., 2005). The expression of PSMA is elevated in other epithelial tumors (de Galiza Barbosa et al., 2020; Lauri et al., 2022), such as tumors in the kidney (Muselaers et al., 2022), breast (Unger et al., 2022), thyroid (Piek et al., 2022), liver (Thompson et al., 2022), pancreas (Sirtl et al., 2021) as well as the lung (Osman et al., 2017). PSMA is expressed in both NSCLC and SCLC (Wang et al., 2015), which suggests a potential clinical utilization in the treatment of lung cancer. We developed software for image segmentation of PSMA areas (Matov and Modiri, 2024) in order to extract morphology metrics regarding the changes after drug treatment (Matov, 2024e) in CTCs to evaluate drug resistance.

Due to the high number of positive PSMA spots per field, the low contrast, and very low signal-to-noise ratio (SNR), the segmentation of PSMA images is technically challenging. Since, the DAPI images have a higher contrast and lower noise in comparison to the PSMA and CD45 channels, we use this channel as a base to process PSMA with a multiscale wavelet method (Olivo-Marin, 2002). Since we expect the same number and position of the PSMA spots as in DAPI images, we use the wavelet segmentation as “seed” areas based on the boundaries found using the DAPI images and improve the boundaries of the DAPI seeds via an active contour procedure using the original PSMA images (Matov and Modiri, 2024). The CD45 images also suffer from low contrast and SNR, similar to the PSMA images. Also, we cannot use the method introduced in segmenting PSMA images, because a DAPI stain is not present in every CD45 position, i.e., not every leukocyte expresses a nuclear marker. For this reason, to segment CD45 images, we have used a damped sine wave (Giancoli, 1988) filtering as a preprocessing step. The combination of the three channels, allows the detection of CTCs as PSMA+/DAPI+/CD45-cells in noisy image datasets, which is the case in particular at low magnification. In high magnification images (20x), the PSMA channel only is sufficient to segment the images and detect CTCs, without having to use the DAPI and CD45 channels (Matov, 2024b; Matov and Modiri, 2024).

To anticipate drug resistance and elucidate mechanisms of resistance, we will derive primary tumor and metastatic organoids from resected tissues from lung cancer patients, and treat them with drug regimens *ex vivo*. Figure 11 shows a lung organoid of about ½ mm in diameter we derived after processing tissue from resected rectal cancer metastasis protruding from the sternum. Patient-derived organoids (Sachs and Clevers, 2014) can be labeled with fluorescent markers of key cellular proteins, which deliver a detailed insight into the metabolic changes in patient tumor cells after drug treatment. We have proposed a computational approach for the selection of drug regimens in solid tumor oncology based on the analysis of EB1 and microtubule dynamics in patient cells (Matov, 2024a; Matov, 2024c; Matov, 2024d; Matov, 2025). Modulation of the microtubule and actin cytoskeleton could sensitize lung cancer cells to immunotherapy. Neoadjuvant anti-PD-1-based immunotherapy might become the standard of care in lung cancer (Topalian and Pardoll, 2025), which further underscores the importance of early detection. A blood-based miR panel has been shown to have a prognostic value for overall survival in advanced stage NSCLC treated with immunotherapy (Rajakumar et al., 2022) and as urinary samples are collected in a non-invasive procedure, prognostic readouts can be obtained more frequently.

The methodology we propose may allow the development of molecular diagnostics tests based on nucleic acid markers for monitoring of cardiovascular, neoplastic, and diseases of the nervous system based on the longitudinal analyses of body fluids. What is novel in this contribution is the description of methodology to detect urine biomarkers for an organ that is not adjacent to the bladder, such as prostate. Further, the approach may provide novel ways for treatment evaluation. Urine voids can be collected at home, the procedure is fully non-invasive, the samples are easy to process (as opposed to blood), the biomarkers are stable in urine, and urine is known to contain disease biomarkers. Correlation between changes in biomarker levels and treatment responses will allow for the early detection of a lack of response after treatment and ultimately for optimal drug selection. It will also facilitate the discovery of biomarkers for the prediction of disease relapse.

In the long run, longitudinal analysis of nucleic acids will allow for the development of novel targeted drugs. Recent literature has described many examples of miRs that could be targeted in disease (Duygu et al., 2016; Kim et al., 2017; Singh and Sen, 2017). Therefore, monitoring of their levels in healthy individuals and patients undergoing disease treatment will likely provide valuable datasets for the pharmaceutical industry as well as for practicing physicians, ultimately allowing them to select the most efficacious treatment sequence and drug combination for each patient. Our overall objective is to significantly improve the quality of life in the last three to four decades of living.

## DECLARATIONS

### Ethics declaration

IRB (IRCM-2019-201, IRB DS-NA-001) of the Institute of Regenerative and Cellular Medicine. Ethical approval was given.

Approval of tissue requests #14-04 and #16-05 to the UCSF Cancer Center Tissue Core and the Genitourinary Oncology Program was given.

### Data availability statement

The datasets used and/or analyzed during the current study are available from the corresponding author upon reasonable request.

## Supporting information

Video 1 - PC3_18_EB1_detections

## Data Availability

All data produced in the present study are available upon reasonable request to the authors. The data to generate the figures is available at: https://github.com/amatov/DiseaseDetectionUrine

https://github.com/amatov/DiseaseDetectionUrine

## ACKNOWLEDGEMENTS

I thank JR&D Services, Rotterdam for funding in the context of NGS data and Neil Bander for the CTC data shown on Fig. 10. I am grateful to the Institute of Regenerative and Cellular Medicine, Santa Monica for issuing the Institutional Review Board protocol approval IRCM-2019-201, IRB DS-NA-001 for the observational study “Longitudinal analysis of next-generation sequencing of nucleic acids for early detection of degenerative diseases such as cardiovascular, neoplastic and diseases related to the nervous system” and James Faber for his feedback regarding the protocol and the process of approval, and to the Genitourinary Tissue Utilization committee and the Genitourinary and Prostate SPORE Tissue Cores at the UCSF Cancer Center for the approval of my tissue requests #14-04 and #16-5 and the Stand Up To Cancer / Prostate Cancer Foundation (SU2C/PCF) West Coast Dream Team (WCDT). The patient blood samples analyzed were from clinical studies with IRB protocols 0804009740 and 0707009283 at Cornell Medicine.

## SUPPLEMENTARY MATERIALS

Video 1 – Time-lapse movie of a cancer organoid with labeled EB1 comets. Organoids obtained from metastasis at a retroperitoneal lymph node of a metastatic castrate-resistant prostate cancer patient. Polymerizing microtubule ends are fluorescently labeled. https://vimeo.com/1057176222/22d94da652

## APPENDIX

### Detection of Alzheimer’s disease (AD)

Evidence for preclinical AD has been identified up to 16 years prior to diagnosis in cerebral fluids (Preische et al., 2019). A list of known blood miRs dysregulated in AD is identified in (Martinez and Peplow, 2019). Of these 42 published AD blood markers, our datasets from NGS sequencing of urine voids shows the presence of 33 miRs (see list below), which is close to 80% of those found in blood plasma, blood serum and cerebrospinal fluid.

### Alzheimer’s disease miRs in blood or cerebrospinal fluid (previously published) and also identified in our urine datasets

miR-146a-5p, miR-106b-3p, miR-195-5p, miR-20b-5p, miR-455-3p, miR-29c-3p, miR-93-5p, miR-19b-3p, miR-501-3p, miR-486-5p, miR-483-5p, miR-502-3p, miR-200a-3p, miR-29a, miR-378a-3p, miR-1291, miR-143-3p, miR-142-3p, miR-328-3p, miR-193a-5p, miR-30a-3p, miR-19b-3p, miR-30d-5p, miR-340-5p, miR-140-5p, miR-125b-5p, miR-26b-5p, miR-16-5p, miR-146a-5p, miR-29a-3p, miR-15b-5p, miR-223-3p (Martinez and Peplow, 2019).

### Detection of Parkinson’s disease (PD)

miR-4639-5p is an example of a marker present in our preliminary urine data which has been already identified in blood (Chen et al., 2017) as a potential biomarker for early diagnosis of PD and also as a potential therapeutic target. Another biomarker detected in our preliminary urine data across all participants is miR-184 (Fig. 12), which has been shown to be negatively regulated by G2019S (Leggio et al., 2017) and other pathogenic LRRK2 mutations (Gehrke et al., 2010), a pathway which we have previously done research studies on in collaboration with Pfizer Inc. (Matov, 2024e).

**Figure 12.**
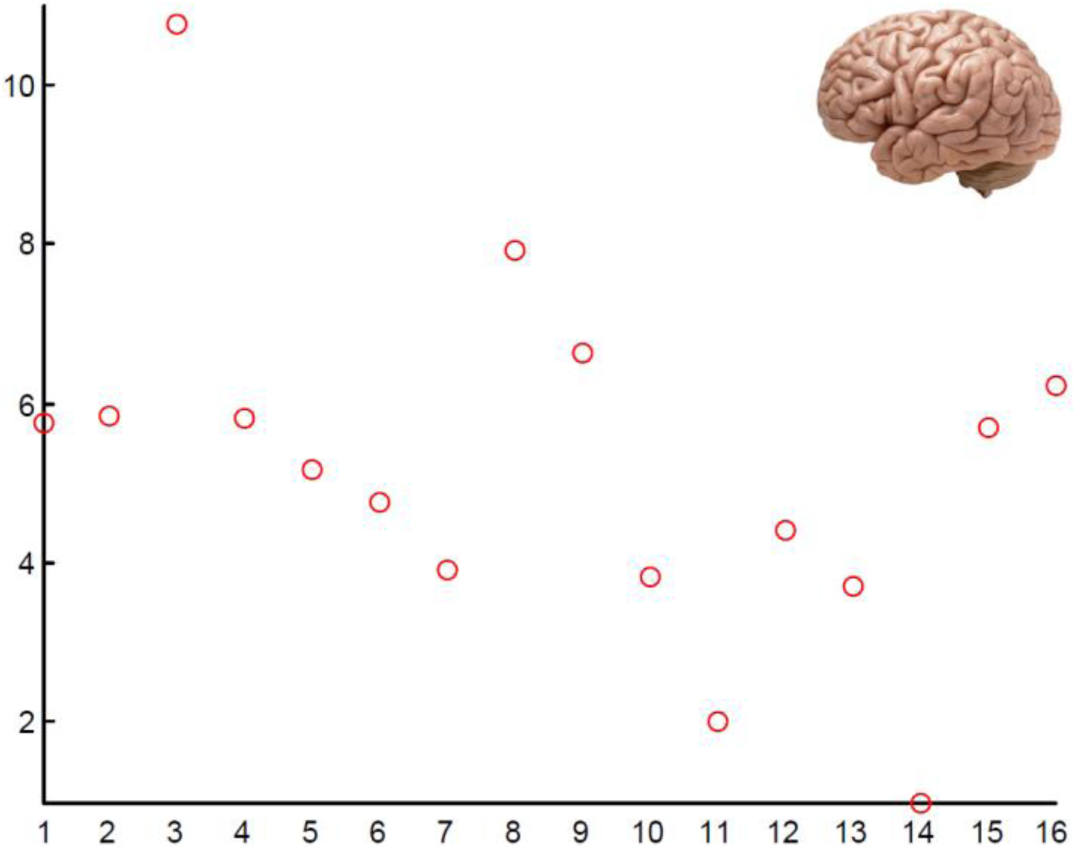
Expression levels (on a logarithmic scale) of miR-184 in urine samples across the 16 patients’ cohort. miR-184 has been shown to negatively regulate G2019S and other pathogenic LRRK2 mutations in Parkinson’s disease. The universal detection among the participants in the cohort demonstrates its potential as a non-invasive urinary biomarker early PD detection test.

### Detection of Huntington’s disease (HD)

Five established plasma markers of HD (Aganzo et al., 2018) are also present in our preliminary data from urine samples and therefore could allow for the development of a noninvasive test for early detection.

### Huntington’s disease miRs in blood (previously published) and also identified in our urine datasets

miR-338-3p, miR-128-3p, miR-23a-3p, miR-24-3p, miR-100-5p (Aganzo et al., 2018).

### Detection of amyotrophic lateral sclerosis (ALS)

25 established cerebrospinal fluid markers of ALS (Ricci et al., 2018) are also present in our preliminary data from urine samples. Therefore, this panel of 25 markers detected could allow for the development of a noninvasive test for early detection.

### Amyotrophic lateral sclerosis miRs in blood (previously published) and also identified in our urine datasets

miR-532-3p, miR-132-3p, miR-143-3p, miR-143-5p, miR-574-5p, miR-338-3p, miR-181a-5p, miR-21-5p, miR-195-5p, miR-15b-5p, miR-let-7a-5p, miR-let-7b-5p, miR-let-7f-5p, miR-124-3p, miR-127-3p, miR-143-3p, miR-125b-2-3p, miR-9-5p, miR-27b-3p, miR-486-5p, miR-16-5p, miR-28-3p, miR-150-5p, miR-378a-3p, miR-142-5p (Ricci et al., 2018).

### Detection of osteoarthritis (OA)

OA is the most common degenerative disease affecting joint tissues. The pathogenesis and progression of OA is poorly understood. miRs play an important role as regulators of underlying biology of cartilage and in pathogenesis (De Palma et al., 2017). Mechanical loading, which is important in the regulation of cartilage metabolism, affects miRs expression. Recent literature suggest that miRs present in human plasma and in synovial fluid could represent promising biomarkers for OA. Six of these established markers are also present in our preliminary data from urine samples and therefore could allow for the development of a noninvasive test for early detection.

### Osteoarthritis miRs in blood (previously published) and also identified in our urine datasets

miR-184, miR-483-5p, miR-27a, miR-199a-5p, miR-199a-3p, miR-582-5p (De Palma et al., 2017).

### Detection of cardiovascular diseases

miRs have been shown to have pervasive roles in cardiovascular biology (Small and Olson, 2011). We have identified in a review article (Li et al., 2013) eight miRs, which are also present in our urine datasets.

### Cardiovascular diseases miRs in blood (previously published) and also identified in our urine datasets

miR-1291 (Li et al., 2013), miR-206 (Lu et al., 2019), miR-15b-5p (Rodriguez and Yin, 2019), miR-122-5p, miR-22-5p (Wang et al., 2019), miR-22-3p (van Boven et al., 2017), miR-423-5p, miR-320b (Gidlof et al., 2013).

### Pancreatic cancer miRs in urine (previously published)

miR-143, miR-223, miR-30e, miR-204 (Debernardi et al., 2015).

### miRs involved in multiple publications on breast cancer (primary tumor site or blood sample; female)

miR-21-5p, miR-9-5p, miR-200a-3p, miR-221-3p, miR-222-3p, miR-155-5p, miR-10b-5p, miR-181a-5p, miR-181b-5p, miR-18a-5p, miR-210-3p, miR-23b-3p, miR-24-3p.

### miRs involved in multiple publications on cancer of the uterus (primary tumor site or blood sample; female)

miR-21-5p, miR-200a-3p, miR-141-3p, miR-200c-3p, miR-200b-3p.

### miRs involved in multiple publications on cervical cancer (primary tumor site or blood sample; female)

miR-21-5p, miR-9-5p, miR-20a-5p, miR-let-7a-5p, miR-10a-5p, miR-146a-5p, miR-205-5p.

### miRs involved in multiple publications on colorectal cancer (primary tumor site or blood sample; female)

miR-21-5p, miR-9-5p, miR-155-5p, miR-27a-3p, miR-let-7a-5p, miR-106b-5p, miR-130a-3p, miR-19a-3p, miR-103a-3p, miR-106a-5p, miR-107, miR-17-5p, miR-196a-5p, miR-23a-3p, miR-200c-3p.

### miRs involved in multiple publications on colorectal cancer (primary tumor site or blood sample; male)

miR-21-5p, miR-9-5p, miR-155-5p, miR-27a-3p, miR-let-7a-5p, miR-106b-5p, miR-130a-3p, miR-19a-3p, miR-103a-3p, miR-106a-5p, miR-107, miR-17-5p, miR-196a-5p, miR-23a-3p, miR-200c-3p.

### miRs involved in multiple publications on prostate cancer (primary tumor site or blood sample; male)

miR-21-5p, miR-221-3p, miR-222-3p, miR-20a-5p, miR-141-3p, miR-125b-5p, miR-148a-3p, miR-182-5p, miR-19a, miR-19b, miR-21.

### miRs involved in multiple (between two and six) publications on lung cancer (primary tumor site or blood sample)

miR-21-3p, miR-21-5p, miR-140-3p, miR-140-5p, miR-155, miR-19b-3p, miR-200b-5p, miR-223, miR-130b-3p, miR-130b-5p, miR-145, miR-200a-3p, miR-200a-5p, miR-205, miR-20a, miR-210, miR-221-3p, miR-339-3p, miR-93, miR-142-3p, miR-142-5p, miR-146a-5p, miR-148a, miR-151-3p, miR-151a-5p, miR-18a-3p, miR-19a, miR-200c, miR-22, miR-26b, miR-28-3p, miR-29a, miR-30a-3p, miR-32, miR-323-5p, miR-340, miR-345, miR-425-5p, miR-4299, miR-454, miR-483-3p, miR-483-5p, miR-766-3p, miR-182, miR-183-3p, miR-197-3p, miR-324-5p, miR-328-3p, miR-424, miR-542-3p, miR-625-3p, miR-625-5p, miR-629, miR-708, miR-9-3p, miR-9-5p.

**miRs involved in lung cancer development, progression and drug resistance** (based on a literature search, which is not exhaustive, i.e., there are more papers on lung cancer with these biomarkers than those listed below):

|  |  |  |
| --- | --- | --- |
| miR-21 | (Wang et al., 2014) | primary |
| miR-21 | (Boeri et al., 2011) | primary |
| miR-21 | (Markou et al., 2016) | primary |
| miR-21 | (Yan et al., 2014) | plasma |
| miR-21 | (Geng et al., 2014) | plasma |
| miR-21-3p | (Nadal et al., 2014) | primary |
| miR-21-5p | (Boeri et al., 2012) | plasma |
| miR-21-5p | (Qu et al., 2017) | plasma, primary |
| miR-140 | (Nadal et al., 2015) | serum |
| miR-140 | (Wozniak et al., 2015) | plasma |
| miR-140-3p | (Nadal et al., 2015) | serum |
| miR-140-5p | (Boeri et al., 2012) | plasma |
| miR-155 | (Wang et al., 2014) | primary |
| miR-155 | (Yan et al., 2014) | plasma |
| miR-155 | (Geng et al., 2014) | plasma |
| miR-155 | (Li et al., 2013) | serum |
| miR-155 | (Wozniak et al., 2015) | plasma |
| miR-19b | (Aushev et al., 2013) | plasma, post-surgery |
| miR-19b | (Wozniak et al., 2015) | plasma |
| miR-19b-3p | (Boeri et al., 2012) | plasma |
| miR-19b-3p | (Zhou et al., 2017) | plasma, primary |
| miR-200b | (Boeri et al., 2011) | primary |
| miR-200b | (Nadal et al., 2015) | serum, primary |
| miR-200b-5p | (Cazzoli et al., 2013) | exosomes |
| miR-200b-5p | (Nadal et al., 2014) | primary |
| miR-223 | (Geng et al., 2014) | plasma |
| miR-223 | (Chen et al., 2012) | serum |
| miR-223 | (Wozniak et al., 2015) | plasma |
| miR-130b | (Wozniak et al., 2015) | plasma |
| miR-130b-3p | (Nadal et al., 2014) | primary |
| miR-130b-5p | (Nadal et al., 2014) | primary |
| miR-145 | (Geng et al., 2014) | plasma |
| miR-145 | (Chen et al., 2012) | serum |
| miR-145 | (Yan et al., 2014) | serum |
| miR-150 | (Markou et al., 2016) | primary |
| miR-150 | (Li et al., 2013) | serum |
| miR-150* | (Leidinger et al., 2016) | plasma, post-surgery |
| miR-200a | (Markou et al., 2016) | primary |
| miR-200a-3p | (Nadal et al., 2014) | primary |
| miR-200a-5p | (Nadal et al., 2014) | primary |
| miR-205 | (Aushev et al., 2013) | plasma , post-surgery |
| miR-205 | (Nadal et al., 2014) | primary |
| miR-205* | (Leidinger et al., 2016) | plasma, post-surgery |
| miR-20a | (Aushev et al., 2013) | plasma , post-surgery |
| miR-20a | (Geng et al., 2014) | plasma |
| miR-20a | (Chen et al., 2012) | serum |
| miR-210 | (Boeri et al., 2011) | primary |
| miR-210 | (Li et al., 2013) | serum |
| miR-210 | (Nadal et al., 2014) | primary |
| miR-221 | (Chen et al., 2012) | serum |
| miR-221-3p | (Boeri et al., 2012) | plasma |
| miR-221-3p | (Zhou et al., 2017) | plasma, primary |
| miR-339-3p | (Leidinger et al., 2014) | blood |
| miR-339-3p | (Nadal et al., 2015) | serum |
| miR-339-3p | (Wozniak et al., 2015) | plasma |
| miR-93 | (Nadal et al., 2015) | serum, primary |
| miR-93 | (Wozniak et al., 2015) | plasma |
| miR-93* | (Nadal et al., 2015) | serum, primary |
| miR-142-3p | (Boeri et al., 2012) | plasma |
| miR-146a | (Yan et al., 2014) | serum |
| miR-146a-5p | (Wozniak et al., 2015) | plasma |
| miR-148 | (Markou et al., 2016) | primary |
| miR-151-3p | (Wozniak et al., 2015) | plasma |
| miR-151a-5p | (Cazzoli et al., 2013) | exosomes |
| miR-18a | (Wozniak et al., 2015) | plasma |
| miR-18a-3p | (Leidinger et al., 2014) | blood |
| miR-19a | (Aushev et al., 2013) | plasma, post-surgery |
| miR-19a | (Nadal et al., 2015) | serum, primary |
| miR-200c | (Nadal et al., 2015) | serum, primary |
| miR-200c | (Wozniak et al., 2015) | plasma |
| miR-22 | (Wozniak et al., 2015) | plasma |
| miR-22* | (Nadal et al., 2015) | serum |
| miR-26b | (Nadal et al., 2015) | serum |
| miR-26b | (Wozniak et al., 2015) | plasma |
| miR-28 | (Wozniak et al., 2015) | plasma |
| miR-28-3p | (Boeri et al., 2012) | plasma |
| miR-29a | (Nadal et al., 2015) | serum |
| miR-30a-3p | (Cazzoli et al., 2013) | exosomes |
| miR-30a-3p | (Wozniak et al., 2015) | plasma |
| miR-32* | (Leidinger et al., 2016) | plasma , post-surgery |
| miR-323 | (Markou et al., 2016) | primary |
| miR-323-5p | (Backes et al., 2016) | serum, pre/post diagnosis |
| miR-340 | (Nadal et al., 2015) | serum |
| miR-340 | (Wozniak et al., 2015) | plasma |
| miR-345 | (Nadal et al., 2015) | serum, primary |
| miR-345 | (Nadal et al., 2014) | primary |
| miR-425 | (Wozniak et al., 2015) | plasma |
| miR-425-5p | (Zhou et al., 2017) | plasma, primary |
| miR-4299 | (Leidinger et al., 2016) | plasma , post-surgery |
| miR-4299 | (Leidinger et al., 2014) | plasma , post-surgery |
| miR-454 | (Wozniak et al., 2015) | plasma |
| miR-454* | (Leidinger et al., 2016) | plasma, post-surgery |
| miR-483-3p | (Leidinger et al., 2016) | plasma, post-surgery |
| miR-483-5p | (Backes et al., 2016) | serum, pre/post diagnosis |
| miR-766 | (Wozniak et al., 2015) | plasma |
| miR-766-3p | (Leidinger et al., 2014) | blood |
| miR-125a-3p | (Leidinger et al., 2016) | plasma, post-surgery |
| miR-125a-5p | (Yan et al., 2014) | serum |
| miR-125b | (Nadal et al., 2015) | serum |
| miR-125b-5p | (Leidinger et al., 2014) | blood |
| miR-1274b | (Leidinger et al., 2016) | plasma , post-surgery |
| miR-1274b | (Leidinger et al., 2014) | blood |
| miR-133b | (Nadal et al., 2015) | serum |
| miR-154-3p | (Cazzoli et al., 2013) | exosomes |
| miR-154-* | (Markou et al., 2016) | primary |
| miR-15b-5p | (Boeri et al., 2012) | plasma |
| miR-17 | (Cazzoli et al., 2013) | exosomes |
| miR-17-5p | (Boeri et al., 2012) | plasma |
| miR-182 | (Li et al., 2013) | serum |
| miR-182 | (Nadal et al., 2014) | primary |
| miR-183 | (Li et al., 2013) | serum |
| miR-183-3p | (Nadal et al., 2014) | primary |
| miR-197 | (Wang et al., 2014) | primary |
| miR-197-3p | (Boeri et al., 2012) | plasma |
| miR-30b | (Aushev et al., 2013) | plasma , post-surgery |
| miR-30b-5p | (Boeri et al., 2012) | plasma |
| miR-30c-5p | (Boeri et al., 2012) | plasma |
| miR-30c-5p | (Leidinger et al., 2014) | blood |
| miR-324-5p | (Boeri et al., 2011) | primary |
| miR-324-5p | (Nadal et al., 2015) | serum |
| miR-328 | (Wang et al., 2014) | primary |
| miR-328-3p | (Leidinger et al., 2014) | blood |
| miR-378a | (Cazzoli et al., 2013) | exosomes |
| miR-378a-5p | (Leidinger et al., 2014) | blood |
| miR-424 | (Markou et al., 2016) | primary |
| miR-424 | (Nadal et al., 2014) | primary |
| miR-451 | (Aushev et al., 2013) | plasma , post-surgery |
| miR-451 | (Backes et al., 2016) | serum, pre/post diagnosis |
| miR-486 | (Li et al., 2013) | serum |
| miR-486-5p | (Leidinger et al., 2016) | plasma, post-surgery |
| miR-542-3p | (Markou et al., 2016) | primary |
| miR-542-3p | (Nadal et al., 2014) | primary |
| miR-574-3p | (Leidinger et al., 2016) | plasma, post-surgery |
| miR-574-5p | (Foss et al., 2011) | serum |
| miR-625-3p | (Nadal et al., 2014) | primary |
| miR-625-5p | (Nadal et al., 2014) | primary |
| miR-629 | (Cazzoli et al., 2013) | exosomes |
| miR-629 | (Nadal et al., 2014) | primary |
| miR-708 | (Wang et al., 2014) | primary |
| miR-708 | (Nadal et al., 2014) | primary |
| miR-720 | (Leidinger et al., 2016) | plasma, post-surgery |
| miR-720 | (Leidinger et al., 2014) | blood |
| miR-885-5p | (Leidinger et al., 2016) | plasma, post-surgery |
| miR-885-5p | (Leidinger et al., 2014) | blood |
| miR-9-3p | (Nadal et al., 2014) | primary |
| miR-9-5p | (Nadal et al., 2014) | primary |
| miR-let-7e* | (Leidinger et al., 2016) | plasma, post-surgery |
| miR-let-7f | (Wozniak et al., 2015) | plasma |
| miR-100 | (Cazzoli et al., 2013) | exosomes |
| miR-101 | (Nadal et al., 2015) | serum |
| miR-106a-5p | (Boeri et al., 2012) | plasma |
| miR-106b | (Nadal et al., 2014) | primary |
| miR-122 | (Backes et al., 2016) | serum, pre/post diagnosis |
| miR-1224-3p | (Leidinger et al., 2016) | plasma, post-surgery |
| miR-1227 | (Leidinger et al., 2016) | plasma, post-surgery |
| miR-1228* | (Backes et al., 2016) | serum |
| miR-1249 | (Leidinger et al., 2016) | plasma, post-surgery |
| miR-1254 | (Foss et al., 2011) | serum |
| miR-126-5p | (Boeri et al., 2012) | plasma |
| miR-1260 | (Leidinger et al., 2016) | plasma, post-surgery |
| miR-1268 | (Foss et al., 2011) | serum |
| miR-1271 | (Wozniak et al., 2015) | plasma |
| miR-1274a | (Leidinger et al., 2014) | blood |
| miR-1280 | (Leidinger et al., 2014) | blood |
| miR-1281 | (Leidinger et al., 2016) | plasma, post-surgery |
| miR-1296 | (Leidinger et al., 2016) | plasma, post-surgery |
| miR-133a | (Nadal et al., 2015) | serum |
| miR-134 | (Leidinger et al., 2014) | plasma , post-surgery |
| miR-135a | (Wang et al., 2014) | primary |
| miR-135b | (Nadal et al., 2014) | primary |
| miR-139-5p | (Cazzoli et al., 2013) | exosomes |
| miR-141 | (Nadal et al., 2015) | serum, primary |
| miR-143 | (Nadal et al., 2015) | serum |
| miR-144 | (Leidinger et al., 2016) | plasma, post-surgery |
| miR-152 | (Chen et al., 2012) | serum |
| miR-181c | (Markou et al., 2016) | primary |
| miR-1825 | (Leidinger et al., 2016) | plasma, post-surgery |
| miR-186 | (Nadal et al., 2015) | serum |
| miR-187* | (Leidinger et al., 2016) | plasma, post-surgery |
| miR-190 | (Markou et al., 2016) | primary |
| miR-190b | (Cazzoli et al., 2013) | exosomes |
| miR-191 | (Wozniak et al., 2015) | plasma |
| miR-1915 | (Leidinger et al., 2014) | plasma , post-surgery |
| miR-193b | (Nadal et al., 2015) | serum, primary |
| miR-195 | (Nadal et al., 2015) | serum |
| miR-196a | (Wang et al., 2014) | primary |
| miR-1974 | (Cazzoli et al., 2013) | exosomes |
| miR-1975 | (Backes et al., 2016) | serum, pre/post diagnosis |
| miR-1976 | (Leidinger et al., 2016) | plasma, post-surgery |
| miR-199a-5p | (Chen et al., 2012) | serum |
| miR-203 | (Markou et al., 2016) | primary |
| miR-204 | (Leidinger et al., 2016) | plasma, post-surgery |
| miR-20b | (Nadal et al., 2015) | serum, primary |
| miR-211 | (Leidinger et al., 2016) | plasma, post-surgery |
| miR-216b | (Nadal et al., 2014) | primary |
| miR-219-1 | (Boeri et al., 2011) | primary |
| miR-222 | (Chen et al., 2012) | serum |
| miR-24 | (Chen et al., 2012) | serum |
| miR-25 | (Chen et al., 2012) | serum |
| miR-27a | (Wang et al., 2014) | primary |
| miR-296 | (Nadal et al., 2015) | serum |
| miR-29c-5p | (Leidinger et al., 2014) | blood |
| miR-301 | (Nadal et al., 2015) | serum |
| miR-30d | (Wozniak et al., 2015) | plasma |
| miR-31 | (Wang et al., 2014) | primary |
| miR-320 | (Chen et al., 2012) | serum |
| miR-329 | (Markou et al., 2016) | primary |
| miR-33 | (Markou et al., 2016) | primary |
| miR-331 | (Nadal et al., 2014) | primary |
| miR-335 | (Wozniak et al., 2015) | plasma |
| miR-346 | (Leidinger et al., 2016) | plasma, post-surgery |
| miR-34b | (Leidinger et al., 2016) | plasma, post-surgery |
| miR-3610 | (Leidinger et al., 2016) | plasma, post-surgery |
| miR-3646 | (Leidinger et al., 2016) | plasma, post-surgery |
| miR-3647-5p | (Leidinger et al., 2016) | plasma, post-surgery |
| miR-371 | (Markou et al., 2016) | primary |
| miR-373 | (Markou et al., 2016) | primary |
| miR-374 | (Nadal et al., 2015) | serum |
| miR-375 | (Nadal et al., 2014) | primary |
| miR-376a-5p | (Cazzoli et al., 2013) | exosomes |
| miR-377 | (Markou et al., 2016) | primary |
| miR-379 | (Cazzoli et al., 2013) | exosomes |
| miR-3922 | (Leidinger et al., 2016) | plasma, post-surgery |
| miR-409-3p | (Zhou et al., 2017) | plasma, primary |
| miR-4254 | (Leidinger et al., 2016) | post-surgery |
| miR-4281 | (Leidinger et al., 2016) | plasma, post-surgery |
| miR-429 | (Nadal et al., 2014) | primary |
| miR-4290 | (Leidinger et al., 2016) | plasma, post-surgery |
| miR-4297 | (Leidinger et al., 2016) | plasma, post-surgery |
| miR-4326 | (Leidinger et al., 2016) | plasma, post-surgery |
| miR-4327 | (Leidinger et al., 2016) | plasma, post-surgery |
| miR-449b* | (Leidinger et al., 2016) | plasma, post-surgery |
| miR-449c* | (Leidinger et al., 2016) | plasma, post-surgery |
| miR-450a | (Nadal et al., 2014) | primary |
| miR-450b | (Nadal et al., 2014) | primary |
| miR-453 | (Markou et al., 2016) | primary |
| miR-484 | (Wozniak et al., 2015) | plasma |
| miR-502-5p | (Cazzoli et al., 2013) | exosomes |
| miR-509 | (Markou et al., 2016) | primary |
| miR-512-5p | (Markou et al., 2016) | primary |
| miR-517b | (Markou et al., 2016) | primary |
| miR-518e | (Markou et al., 2016) | primary |
| miR-520e | (Nadal et al., 2014) | primary |
| miR-583 | (Leidinger et al., 2016) | serum, pre/post diagnosis |
| miR-584-5p | (Zhou et al., 2017) | plasma |
| miR-590-3p | (Wozniak et al., 2015) | plasma |
| miR-595 | (Backes et al., 2016) | serum, pre/post diagnosis |
| miR-605 | (Leidinger et al., 2016) | plasma , post-surgery |
| miR-609 | (Leidinger et al., 2016) | plasma , post-surgery |
| miR-613 | (Leidinger et al., 2016) | plasma, post-surgery |
| miR-634 | (Leidinger et al., 2016) | plasma, post-surgery |
| miR-647 | (Leidinger et al., 2016) | plasma, post-surgery |
| miR-650 | (Nadal et al., 2014) | primary |
| miR-652 | (Wozniak et al., 2015) | plasma |
| miR-663 | (Wang et al., 2014) | primary |
| miR-7-2-prec | (Boeri et al., 2011) | primary |
| miR-744 | (Nadal et al., 2015) | serum, primary |
| miR-758 | (Nadal et al., 2015) | serum, primary |
| miR-762 | (Leidinger et al., 2016) | plasma , post-surgery |
| miR-764 | (Leidinger et al., 2016) | plasma, post-surgery |
| miR-767-3p | (Backes et al., 2016) | serum, pre/post diagnosis |
| miR-889 | (Nadal et al., 2014) | primary |
| miR-937 | (Leidinger et al., 2016) | plasma, post-surgery |
| miR-941 | (Leidinger et al., 2016) | plasma, post-surgery |
| miR-942-5p | (Leidinger et al., 2014) | blood |
| miR-96 | (Markou et al., 2016) | primary |
| miR-99b* | (Backes et al., 2016) | serum, pre/post diagnosis |

